# Identification of Major Congenital Malformations based on Healthcare Databases in France: a Proof-of-Concept Study using the EPI-MERES Nationwide Mother-Child Register

**DOI:** 10.64898/2026.03.06.26347780

**Authors:** Tom Duchemin, Lise Marty, Jérôme Drouin, Sara Miranda, Jérémie Botton, Valérie Olié, Alain Weill, Rosemary Dray-Spira

## Abstract

**Aim:** Besides registries, healthcare databases can provide useful information for assessing major congenital malformations (MCMs) frequency and investigating their risk factors, particularly medications exposures. This study aimed to assess the validity of MCMs identification based on French national, comprehensive healthcare databases.

**Methods:** Using information on hospital discharge diagnoses, medical procedures (e.g. surgery) and death causes from the EPI-MERES register nested in the French National Health Data System, 72 specific MCMs grouped in 11 organ groups were assessed among all births occurred after 22 weeks of amenorrhea in France between 2010 and 2023. MCMs prevalence rates were estimated and compared to those from EUROCAT, and associations with prenatal exposure to valproate were assessed.

**Results:** Among 10.5 million births, 213,153 live born infants with at least one MCM, i.e. 203.0 cases per 10,000 births, were identified. MCMs prevalence rates among live births were close to those reported in EUROCAT overall (difference: -1.76 per 10,000 births [-1%]), for each organ group (differences ranging from -9.10 [-13%] to +3.44 [+16%] per 10,000 births), and for the 72 specific MCMs (median prevalence difference: 1%). Prenatal exposure to valproate was significantly associated with increased risks of any MCM (adjusted odds ratio (aOR) 2.82, 95% CI [2.33–3.41]) and of 15 specific MCMs including spina bifida (aOR 17.88 [7.88–40.53]).

**Conclusion:** This study supports the validity of MCMs identification based on data of the EPI-MERES register. The EPI-MERES register provides a highly powerful, reactive and operational tool complementing MCMs registries for improving real-life knowledge on drug teratogenicity.

**Plain language summary:** Major congenital malformations are serious structural abnormalities present at birth that can have lasting consequences on children’s health. Better understanding their risk factors, particularly medication exposures during pregnancy, is crucial. Population-based registries are today the primary source of information on malformations, but healthcare databases could offer a faster and broader alternative. This study tested whether the EPI-MERES register, built upon the French National Health Data System (SNDS), could accurately identify 72 specific malformations across 10.5 million births between 2010 and 2023. Prevalence estimates closely matched those from the European EUROCAT registry, confirming good data accuracy. As expected, valproate (a known teratogen) was associated with an increased risk of various malformations, including spina bifida, EPI-MERES thus constitutes a promising tool for studying medication risks during pregnancy.

## Introduction

Major Congenital Malformations (MCM) are defined by the World Health Organization as structural anomalies developing in utero which have a significant medical, social or aesthetic impact for the individual ^1^. In Europe, according to the European network for the surveillance of congenital anomalies EUROCAT MCMs concern 2.6% of all births and 2.0% of live births ^2^. They account for 27.7% of deaths among children under five years of age in high-income countries ^3^ and otherwise may induce life-long disability. MCMs can result from a range of factors such as *in utero* exposure to infectious agents, alcohol, tobacco or environmental toxic substances ^1^. Prenatal exposure to certain medications is also a cause of MCMs, as evidenced, for example, by the well-documented teratogenic effects of valproate—an antiepileptic drug—reported in numerous studies ^4,5^.

In Europe, one of the primary sources of information on MCMs is EUROCAT, a collaborative European network of 44 population-based congenital anomalies registries in 23 European countries covering approximately 1.5 million births per year (https://eu-rd-platform.jrc.ec.europa.eu/eurocat_en). EUROCAT provides high-quality epidemiological information for research on MCMs causes, prevention, treatment and care ^2^. However, while registry-based data offer high-reliability and valuable insights, they also come with limitations including high operational costs, delayed data availability, and incomplete geographical coverage. These constraints can affect both the timeliness and representativeness of the data. Additionally, MCMs registries often collect only limited information on prenatal exposures (e.g., medication exposures) potentially increasing the risks of birth defects. Furthermore, they generally do not include data on control populations, such as births with no MCM, limiting comparative analyses.

Besides registries, healthcare databases have the potential to provide at low cost routinely collected data of interest for assessing MCMs frequency and investigating and their risk factors, particularly medications exposures, among large populations and over long periods. However, healthcare databases are not primarily designed for epidemiological research, which may cause many challenges to properly identify MCMs. Previous reports based on such databases showed substantial differences in MCMs assessment depending on data sources ^6^, targeted MCMs ^7,8^ and length of follow-up after birth ^9^. While some studies suggest a high accuracy of MCMs identification in healthcare databases ^8,10^, validity studies remain essential to assess the performance of algorithms used for case identification.

This study aimed to assess the validity of MCMs identification based on French national, comprehensive healthcare databases. Using data of the EPI-MERES register nested in the French National Health Data System (SNDS), MCMs prevalence was assessed among children born in France between 2010 and 2023, and compared to estimates from EUROCAT. Furthermore, as a proof-of-concept analysis, the association between prenatal exposure to valproate - a drug with known teratogenic effects - and MCMs was assessed.

## Methods

### Data source

The National Mother-Child EPI-MERES Register includes all pregnancies of women aged 12-55 that ended in France since January 1, 2010 and their related offsprings, if any. It is built upon data of the French National Health Data System (Système National des Données de Santé, SNDS) ^11^, which covers approximately 67 million people (more than 99% of the French population) and is updated annually, with data of a given year available by July of the following year. Data of the offspring are linked to those of their mother for 97% of all births, and for more than 99% of live born infants. The EPI-MERES Register has been previously used for several studies ^12–17^

EPI-MERES includes comprehensive individual information on pregnancies (pregnancy start and end dates, pregnancy course and outcome, medications reimbursed during pregnancy with corresponding dispensation dates) and on mothers and offsprings (sociodemographic characteristics, reimbursed outpatient care and medications, hospital discharge diagnoses [coded using FR-ICD-10 classification] and medical procedures including surgical, medical and imaging acts [coded using the French CCAM classification], date and cause of death [coded using FR-ICD-10 classification] if appropriate).

### Study Population

We included all births (live births, stillbirths and pregnancy terminations) with available mother-offspring linkage occurred after 22 weeks of amenorrhea (foetal viability threshold) between January 1, 2010 and December 31, 2023.

### MCM identification

Based on the EUROCAT list included in Guide 1.5 ^18^, 72 specific MCMs (listed in **Table 1**) were identified using information on hospital discharge diagnoses, death causes from death certificates (if appropriate) and, for 22 of these 72 MCMs, specific medical procedures. For MCMs whose identification required a medical procedure (e.g. malformation correction surgery), information from hospital discharge diagnoses and/or death certificates were deemed sufficient when death occurred by the age of 6 months or when a death certificate included a FR-ICD-10 code of MCM. FR-ICD-10 codes used for MCMs identification were similar to those used in EUROCAT, except for MCMs identified by four-digit ICD-10 codes for which identification criteria were adapted given that only three-digit codes are available in the SNDS. Differences between EUROCAT and EPI-MERES codes lists are detailed in **Table S1.**

**Table 1.** Major Congenital Malformations identified in the EPI-MERES register.

|  |  |
| --- | --- |
| **Anomalies of the nervous system** | Anencephaly and similar malformations ; Encephalocele ; Spina Bifida ; Congenital hydrocephalus ; Microcephaly ; Arhinencephaly/Holoprosencephaly ; Congenital malformations of corpus callosum. |
| **Anomalies of the eyes** | Cystic eyeball/Other anophthalmos/Microphthalmos ; Congenital cataract ; Congenital glaucoma. |
| **Anomalies of the ear, face and neck** | Congenital absence of (ear) auricle/Congenital absence atresia and structure of auditory canal (external). |
| **Congenital heart defects** | Common arterial trunk ; Double outlet right ventricle ; Double outlet left ventricle ; Discordant ventriculoarterial connection ; Discordant atrioventricular connection ; Double inlet ventricle ; Ventricular septal defect ; Atrial septal defect, incl. persistent foramen ovale ; Atrioventricular septal defect ; Tetralogy of Fallot ; Congenital tricuspid stenosis ; Ebstein anomaly ; Congenital pulmonary valve stenosis ; Pulmonary valve atresia ; Congenital stenosis of aortic valve ; Congenital mitral stenosis ; Hypoplastic left heart syndrome ; Hypoplastic right heart syndrome ; Coarctation of aorta ; Atresia of aorta ; Total anomalous pulmonary venous connection ; Patent ductus arteriosus. |
| **Respiratory anomalies** | Choanal atresia. |
| **Oro-facial clefts** | Cleft palate ; Cleft lip/Cleft palate with cleft lip. |
| **Anomalies of the digestive system** | Atresia of oesophagus with/without tracheo-oesophageal fistula ; Congenital absence, atresia and stenosis of duodenum ; Congenital absence, atresia and stenosis of jejunum/ileum/ other specified parts of small intestine ; Congenital absence, atresia and stenosis of anus/rectum with/without fistula ; Hirschprung disease ; Congenital malformations of intestinal fixation ; Atresia of bile ducts ; Annular pancreas ; Congenital diaphragmatic hernia. |
| **Abdominal wall defects** | Gastroschisis ; Exomphalos. |
| **Congenital anomalies of kidney and urinary tract** | Unilateral renal agenesis ; Bilateral renal agenesis/Potter syndrome ; Renal dysplasia ; Congenital hydronephrosis/Atresia and stenosis of ureter/Other obstructive defects of renal pelvis and ureter ; Lobulated, fused and horseshoe kidney/Ectopic kidney ; Epispadias/Exstrophy of urinary bladder ; Congenital posterior urethral valves ; Prune belly syndrome. |
| **Genital** | Hypospadias ; Indeterminate sex and pseudohermaphroditism. |
| **Limb anomalies** | Reduction defects of upper/lower/unspecified limb ; Talipes equinovarus ; Congenital dislocation of hip, unilateral/bilateral/unspecified ; Polydactyly ; Syndactyly. |
| **Other anomalies** | Craniosynostosis ; Situs inversus ; Septo-optic dysplasia ; Laterality anomalies. |
| **Chromosomal** | Skeletal dysplasia ; Down syndrome ; Trisomy 13/Patau syndrome ; Trisomy 18/Edwards syndrome ; Turner syndrome ; Triploidy and polyploidy. |

In line with EUROCAT, MCMs were grouped together using two aggregation levels: 1) all MCMs combined (any MCM) and 2) categorization into 13 groups: 11 organ groups, one category grouping chromosomal and genetic anomalies, and one including other, unclassified anomalies. In EUROCAT, MCMs groups include both specific listed MCMs and additional MCMs identified with ICD-10 codes corresponding to additional specific, “unspecified” or “other” malformations. However, given that in EPI-MERES MCMs coded as “unspecified” or “other” may refer to minor congenital anomalies, only malformations identified by a specific FR-ICD-10 code were considered for the grouping. In addition, MCMs coded as “Congenital malformation syndromes due to known exogenous causes, not elsewhere classified” (ICD-10 code Q86) or as “Other specified congenital malformation syndromes affecting multiple systems” (ICD-10 code Q87) were included in the “Other anomalies” category rather than in the group “Chromosomal and genetic anomalies” as in EUROCAT, given that ICD-10 codes Q86 and Q87 do not mention a chromosomal or genetic cause. A full description of the 72 specific MCMs and of the 13 groups’ identification criteria is available in supplemental material (**Table S2**).

Since MCMs are not always diagnosed or recorded right after birth, MCMs identification was based on diagnoses and procedures occurring up to 12 months after birth among infants born alive - except for microcephaly, hypospadias, and epispadia for which a time period of 24 months after birth was considered because diagnoses and related medical procedures often take place between 1 and 2 years of age. At the time of the analyses, data on hospital discharge diagnoses and procedures were available until December 31, 2024 and data on death certificates were available until December 31, 2022.

As same-sex twins cannot be distinguished in hospital databases, MCMs identified based on ICD-10 codes only were arbitrarily attributed to the twin with the highest number of healthcare reimbursement claims in the two years following birth.

### MCMs prevalence rates estimation in EPI-MERES and comparison with EUROCAT

All MCMs were assessed among live born infants. In addition, the 11 following MCMs, which are detectable prenatally and severe enough to justify pregnancy termination ^19^, were assessed among stillbirths and foetuses deriving from pregnancies terminated after 22 weeks: spina bifida, bilateral renal agenesis, anencephaly, encephalocele, congenital hydrocephalus, reduction defects of upper/lower/unspecified limb, discordant ventriculoarterial connection, hypoplastic left heart syndrome, congenital diaphragmatic hernia, omphalocele, and gastroschisis.

In line with EUROCAT definition, prevalence rates among live births were estimated as the number of live births with MCM per 10,000 live- and stillbirths. For the 11 MCMs also assessed for stillbirths and interrupted pregnancies, prevalence rates were estimated as the number of live births, stillbirths and pregnancy terminations per 10,000 live- and stillbirths. Time trends in MCMs prevalence rates were assessed across five sub periods: 2010-2012, 2013-2015, 2016-2018, 2019-2021 and 2022-2023. MCMs prevalence rates estimated in EPI-MERES between 2010 and 2023 were compared to the updated EUROCAT estimates available at https://eu-rd-platform.jrc.ec.europa.eu/eurocat/eurocat-data/prevalence_en. Since EUROCAT data for year 2023 were not yet available at the time of the analyses (March 2025), estimates were computed for the period 2010-2022. Absolute and relative differences in prevalence estimates from EPI-MERES and EUROCAT were computed for each individual MCM.

### Associations with prenatal valproate exposure

Associations between valproate monotherapy exposure during the first two months of pregnancy and MCMs occurrence were assessed in live born infants. Given the major changes occurred in prenatal exposure to valproate after 2015 concomitantly with risk reduction measures reinforcement in France ^20^, analyses were restricted to the period 2010-2015. All children born alive between 2010 and 2015 were included, except those prenatally exposed to a teratogenic drug (**Table S3**) or a teratogenic infection (**Table S4**), and those with a chromosomal or genetic anomaly. Prenatal exposure to valproate monotherapy was defined as one or more reimbursements for valproic acid (ATC code N03AG01, excluding divalproic acid), but not for any other antiepileptic drug, between 1 month before and two months after conception. The comparison group included births unexposed to any antiepileptic drug between 1 month before and two months after conception.

Adjusted odds-ratios (aORs) of MCMs associated with prenatal exposure to valproate monotherapy were computed using propensity score-based standardized mortality ratio (SMR) weighted logistic regression models to target an average treatment effect among the treated (ATT) population. The propensity score was estimated using a logistic regression model including socio-demographic characteristics, comorbidities, lifestyle factors and comedications. Generalized estimating equations were used to account for multiple births from a same mother in the study period. All codes used to define exposure and covariates are described in **Table S5**. The robustness of the results was assessed in sensitivity analyses considering two alternative definitions of the outcome: i) live birth with any MCM as identified using FR-ICD-10 codes corresponding to “other unspecified” malformations in addition to specific codes; and ii) composite outcome combining live birth with any MCM, stillbirth or pregnancy termination after 22 weeks of amenorrhea. Analyses were performed with SAS® software version 9.4 (SAS Institute) and with R version 4.3.3 .

## Results

### Study population

Among a total of 14,110,517 pregnancies ended between 2010 and 2023 in the EPI-MERES register, 10,456,252 ended with a pregnancy termination after 22 weeks of amenorrhea, stillbirth or live birth. After exclusion of 101,138 pregnancies with missing mother-infant/foetus linkage, 10,355,114 pregnancies were included. These pregnancies concerned 6,535,333 distinct mothers and ended in 10,529,062 infants/foetuses, out of which 10,458,192 (99.3%) were live births, 41,596 (0.4%) stillbirths and 29,274 (0.3%) pregnancy terminations.

### MCMs prevalence in EPI-MERES

#### Among live births

Overall, a total of 213,153 live born infants with at least one MCM were identified, corresponding to a prevalence rate of 203.0 cases per 10,000 births. This rate increased by 16% between 2010-2012 and 2022-2023, from 181.2 to 211.2 cases per 10,000 births (**Figure 1** and **Table S6**).

**Figure 1.**
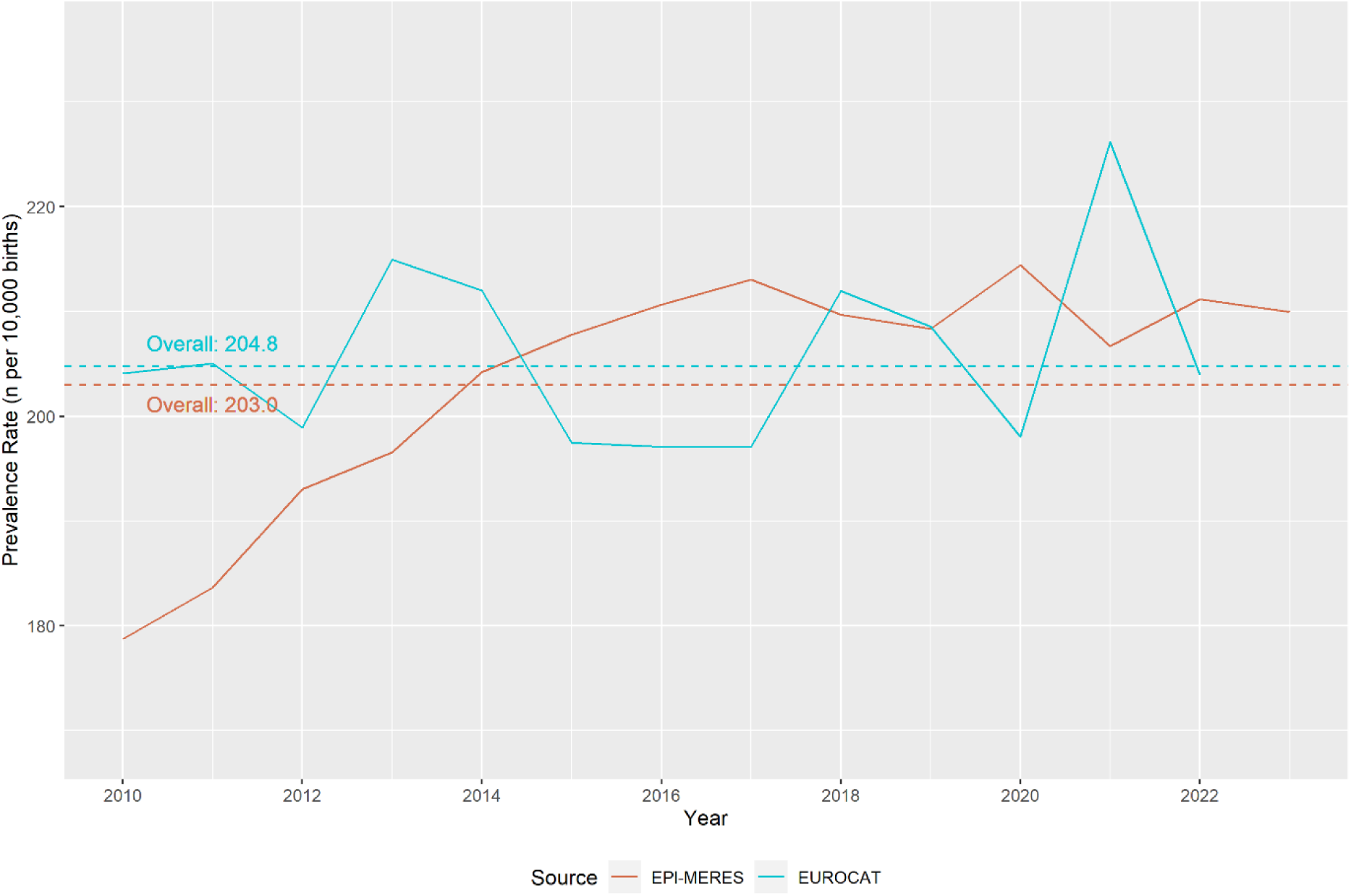
Prevalence of any MCM in EPI-MERES (between 2010 and 2023) and EUROCAT (between 2010 and 2022), overall and by year of birth.

Congenital heart defects accounted for the most prevalent group of MCMs (62.3 cases per 10,000 births), followed by anomalies of the kidney and urinary tract (32.4 per 10,000), limb anomalies (30.4 per 10,000), genital anomalies (24.4 per 10,000), gastrointestinal anomalies (13.9 per 10,000), genetic and chromosomal anomalies (12.9 per 10,000), oro-facial clefts (12.5 per 10,000), nervous system anomalies (11.2 per 10,000), eye anomalies (4.6 per 10,000), respiratory anomalies (3.4 per 10,000), abdominal wall defects (2.8 per 10,000), and ear, face, and neck anomalies (0.7 per 10,000) (**Figure 2**). Prevalence rates of the various MCMs groups showed contrasting trends over time between 2010-2012 and 2022-2023, with consistent increases for nervous system anomalies (+5.0 per 10,000, i.e. +56%), genetic and chromosomal anomalies (+4.5 per 10,000, i.e. +42%), congenital heart defects (+16.6 per 10,000, i.e. +33%), gastrointestinal anomalies (+3.6 per 10,000, i.e. +29%), respiratory anomalies (+0.8 per 10,000, i.e. +28%) and limb anomalies (+6.1 per 10,000, i.e. +23%), consistent decreases for genital anomalies (-2.2 per 10,000, i.e. -9%) and oro-facial clefts (-0.7 per 10,000, i.e. - 5%), and mixed trends for the other MCMs groups (**Figure S1** and **Table S6**). Nine specific MCMs had a prevalence rate higher than 5 cases per 10,000 births among live born infants (**Figure 3**): ventricular septal defect (29.4 cases per 10,000 births), atrial septal defect (including foramen ovale) (24.1 per 10,000), cleft palate (4.3 per 10,000), congenital hydronephrosis (14.8 per 10,000), hypospadias (21.8 per 10,000), club foot/talipes equinovarus (9.0 per 10,000), hip dislocation (6.7 per 10,000), polydactyly (11.0 per 10,000) and Down syndrome (6.3 per 10,000). Detailed prevalence rates of all specific MCMs, overall and by sub periods, are shown in **Table S6**.

**Figure 2.**
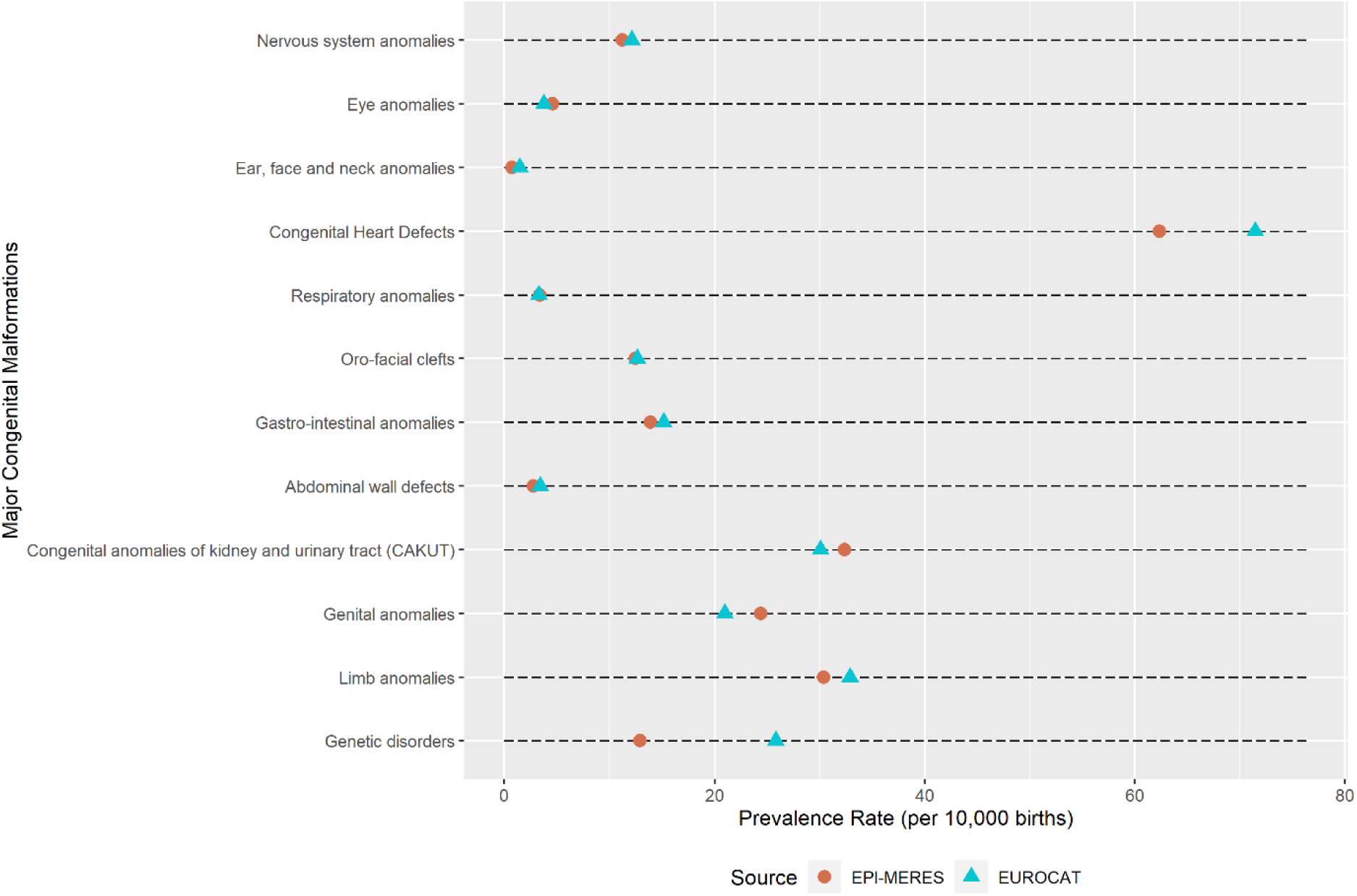
Prevalence of MCMs by organ groups in EPI-MERES (2010 – 2023) and in EUROCAT (2010 – 2022)

**Figure 3.**
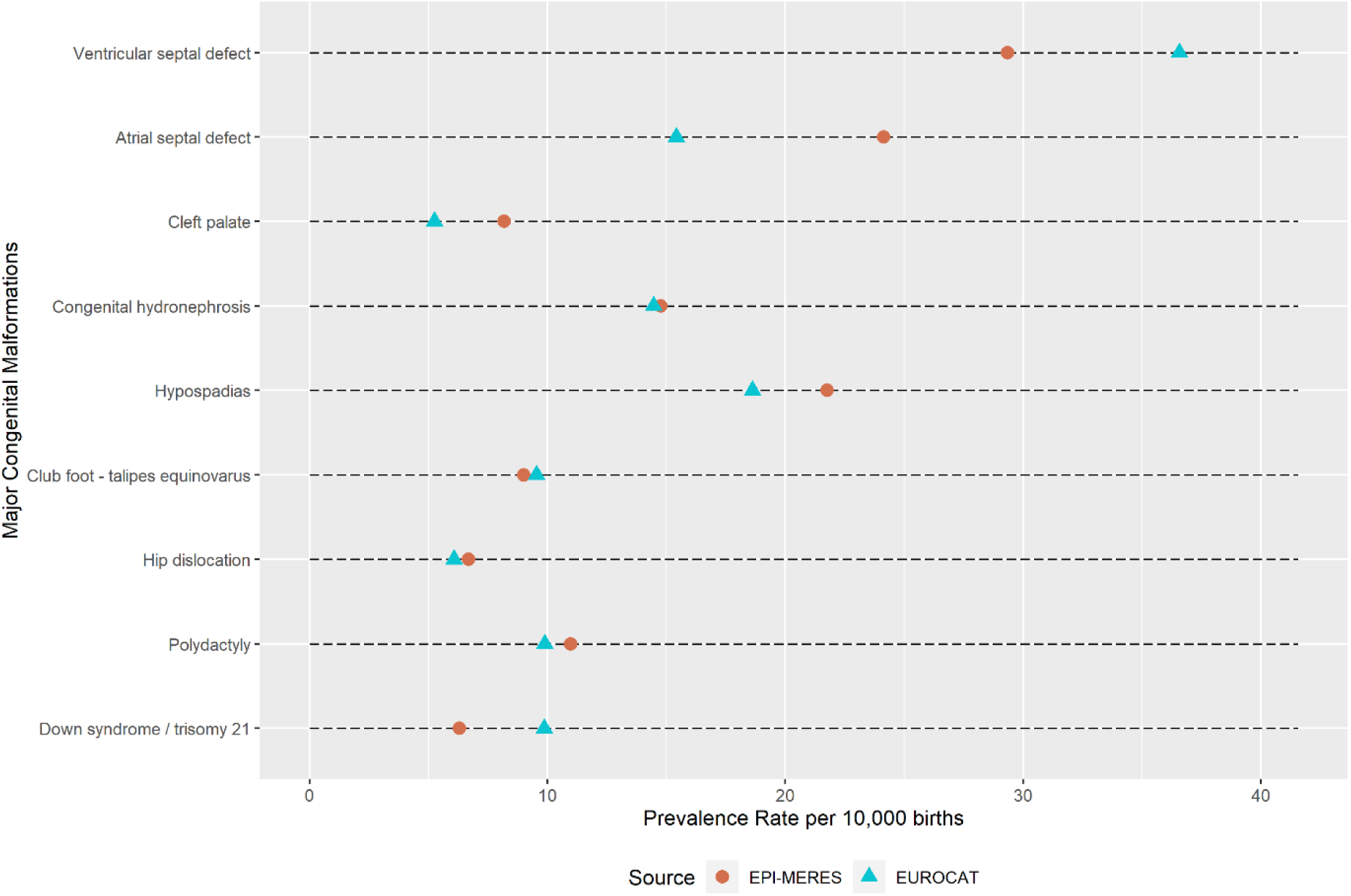
Prevalence of the MCM with a prevalence higher than 5 per 10,000 live births in EPI-MERES and EUROCAT.

#### Among live births, stillbirths and pregnancy terminations

Prevalence rates per 10,000 births among live births, stillbirths and pregnancy terminations reached 0.3 for anencephaly and similar malformations, 0.6 for encephalocele, 2.8 for spina bifida, 3.0 for congenital hydrocephalus, 3.2 for discordant ventriculoarterial connection, 1.7 for hypoplastic left heart syndrome, 2.4 for congenital diaphragmatic hernia, 1.4 for gastroschisis, 1.6 for exomphalos, 0.5 for bilateral renal agenesis/Potter syndrome, 2.4 for reduction defects of upper/lower/unspecified limb (**Table S7**).

### Comparison with EUROCAT

MCMs prevalence rates estimates among live births in EPI-MERES were close to those reported in EUROCAT overall (difference in prevalence rates of any MCM in EPI-MERES *versus* EUROCAT: -1.8 per 10,000 births, i.e. -1%), for the 11 MCMs organ groups (prevalence differences ranging from -9.1 per 10,000 births (-13%) for congenital heart defects to +3.4 per 10,000 births (+16%) for genital anomalies), and for the 72 specific MCMs (median prevalence difference: 1%; interquartile range: -23% to +27%). The prevalence difference was more marked for the group of chromosomal and genetic anomalies: -12.8 per 10,000 births (-50%). For the nine most prevalent specific MCMs, prevalence differences ranged from -7.3 per 10,000 births (-20%) for ventricular septal defect to +8.8 per 10,000 births (+58%) for atrial septal defect (**Figures 2 and 3, Table S6**).

In contrast, MCMs prevalence rates estimates among live births, stillbirths and pregnancy terminations were substantially lower in EPI-MERES compared to EUROCAT, with prevalence differences for the 11 MCMs assessed among live births, stillbirths and pregnancy terminations reaching -46% in median (interquartile range: -59% to -40%) (**Table S7**).

### Associations with prenatal valproate exposure

Among children born alive between 2010 and 2015, 2,091 exposed to valproate monotherapy and 4,683,744 unexposed were included (**Figure S2**). Compared to unexposed children, those exposed were born from mothers who were older, more socially disadvantaged and had more comorbidities (**Table S8**). After propensity score weighting, characteristics were well balanced between the two groups (**Figure S3**).

The prevalence of any MCM was significantly higher in the exposed than in the unexposed group (530.8 versus 181.2 cases per 10,000 live births; aOR 2.8, 95% CI [2.3–3.4]). Significant associations were observed for 7 of the 11 MCMs organ groups: nervous system anomalies (aOR 4.3 [2.3-8.1]), anomalies of the ear, face and neck (aOR 13.8 [3.4-4.4]), congenital heart defects (aOR 3.2 [2.3-4.4]), oro-facial clefts (aOR 3.59 [1.9–6.6]), anomalies of the digestive system (aOR 2.5 [1.2-5.2]), genital anomalies (aOR 3.7 [2.4-5.9]) and limb anomalies (aOR 1.9 [1.1-3.4]). Fifteen of the sixty-eight studied specific MCMs were associated with prenatal exposure to valproate: spina bifida (aOR 17.9 [7.9–40.5]), congenital absence of auricle/congenital absence atresia and structure of auditory canal (aOR 17.6 [4.3–71.5]), ventricular septal defect (aOR 3.3 [2.1-5.10), atrial septal defect (aOR 4.36 [2.8–6.7]), tetralogy of Fallot (aOR 4.2 [1.3-13.0]) pulmonary valve atresia (aOR 8.5 [2.1–34.3], congenital stenosis of aortic valve (aOR 7.3 [1.8–29.6]), hypoplastic left heart syndrome (aOR 11.9 [3.8–37.3]), choanal atresia (aOR 9.8 [1.4–70.9]), cleft palate (aOR 6.2 [2.8–13.9]), congenital malformations of intestinal fixation (aOR 15.5 [4.9–48.5]), hypospadias (aOR 4.1 [2.6-6.5]), talipes equinovarus (aOR 2.6 [1.1–6.2]) polydactyly (aOR 3.0 [1.3–6.7]) and craniosynostosis (aOR 13.2 [7.2-24.0]) (**Table 2**). The association between valproate exposure and any MCM remained unchanged in sensitivity analyses (**Table S9**).

**Table 2.** Association between Valproate Exposure and Major Congenital Malformations in Live births (2010 - 2015)

|  | Exposed to valproate<br>n (n per 10,000 live births) | Non-exposed to AE drugs<br>n (n per 10,000 live births) | crude OR [95%-CI] | adjusted OR [95%-CI] |
| --- | --- | --- | --- | --- |
| **All anomalies** | **111 (530.9)** | **84,865 (181.2)** | 3.0 [2.5 - 3.7] | **2.82 [2.33 - 3.41]** |
| **Anomalies of the nervous system** | **10 (47.8)** | **4,376 (9.3)** | 5.1 [2.8 - 9.6] | **4.33 [2.32 - 8.07]** |
| Anencephaly and similar malformations | 0 (0) | 39 (0.08) | / | / |
| Encephalocele | 0 (0) | 188 (0.4) | / | / |
| Spina Bifida | 6 (28.7) | 645 (1.4) | 21.0 [9.3 - 46.7] | 17.88 [7.88 - 40.53] |
| Congenital hydrocephalus | 1 (4.8) | 1,181 (2.5) | 1.9 [0.3 - 13.5] | 1.59 [0.22 - 11.3] |
| Microcephaly | 2 (9.6) | 1,364 (2.9) | 3.3 [0.8 - 13.2] | 2.58 [0.64 - 10.36] |
| Arhinencephaly/Holoprosencephaly | 0 (0) | 60 (0.1) | / | / |
| Congenital malformations of corpus callosum | 1 (4.8) | 889 (1.9) | 2.5 [0.4 - 17.9] | 2.2 [0.31 - 15.68] |
| **Anomalies of the eyes** | **2 (9.6)** | **2,003 (4.3)** | **2.2 [0.6 – 9.0]** | **2.02[0.5 - 8.1]** |
| Cystic eyeball/Other<br>anophthalmos/Microphthalmos | 1 (4.8) | 328 (0.7) | 6.8 [1.0 - 48.7] | 5.8 [0.8 - 41.7] |
| Congenital cataract | 0 (0) | 630 (1.4) | / | / |
| Congenital glaucoma | 0 (0) | 438 (0.9) | / | / |
| **Anomalies of the ear, face and neck** | **2 (9.6)** | **291 (0.6)** | **15.4 [3.8 – 62.0]** | **13.8 [3.4 - 55.9]** |
| Congenital absence of (ear) auricle/Congenital<br>absence atresia and structure of auditory canal<br>(external) | 2 (9.6) | 228 (0.5) | 19.7 [5.0 - 79.2] | 17.6 [4.3 - 71.5] |
| **Congenital heart defects** | **40 (191.3)** | **24,566 (52.5)** | **3.7 [2.7 - 5.1]** | **3.2 [2.4 - 4.4]** |
| Common arterial trunk | 0 (0) | 182 (0.4) | / | / |
| Double outlet right ventricle | 1 (4.8) | 428 (0.9) | 5.2 [0.7 - 37.3] | 4.6 [0.7 – 33.0] |
| Double outlet left ventricle | 0 (0) | 34 (0.07) | / | / |
| Discordant ventriculoarterial connection | 0 (0) | 1,457 (3.1) | / | / |
| Discordant atrioventricular connection | 0 (0) | 55 (0.1) | / | / |
| Double inlet ventricle | 0 (0) | 269 (0.6) | / | / |
| Ventricular septal defect | 20 (95.7) | 12,188 (26.0) | 3.7 [2.4 - 5.8] | 3.3 [2.1 - 5.1] |
| Atrial septal defect, incl. persistent foramen ovale | 21 (100.3) | 8,496 (18.1) | 5.6 [3.6 - 8.6] | 4.5 [2.8 - 6.7] |
| Atrioventricular septal defect | 1 (4.8) | 794 (1.7) | 2.8 [0.4 - 20.1] | 2.5 [0.3 - 17.5] |
| Tetralogy of Fallot | 3 (14.4) | 1,425 (3.0) | 4.7 [1.5 - 14.7] | 4.2 [1.3 - 13.0] |
| Congenital tricuspid stenosis | 0 (0) | 186 (0.4) | / | / |
| Ebstein anomaly | 0 (0) | 132 (0.3) | / | / |
| Congenital pulmonary valve stenosis | 2 (9.6) | 1,378 (2.9) | 3.3 [0.8 - 13.0] | 2.9 [0.7 - 11.6] |
| Pulmonary valve atresia | 2 (9.6) | 466 (1.0) | 9.6 [2.4 - 38.6] | 8.5 [2.1 - 34.3] |
| Congenital stenosis of aortic valve | 2 (9.6) | 489 (1.0) | 9.2 [2.3 - 36.8] | 7.3 [1.8 - 29.6] |
| Congenital mitral stenosis | 0 (0) | 87 (0.2) | / | / |
| Hypoplastic left heart syndrome | 3 (14.4) | 538 (1.2) | 12.5 [4.0 - 38.9] | 11.9 [3.8 - 37.3] |
| Hypoplastic right heart syndrome | 0 (0) | 189 (0.4) | / | / |
| Coarctation of aorta | 2 (9.6) | 1,581 (3.4) | 2.8 [0.7 - 11.4] | 2.7 [0.7 - 11.0] |
| Atresia of aorta | 0 (0) | 163 (0.4) | / | / |
| Total anomalous pulmonary venous connection | 0 (0) | 314 (0.7) | / | / |
| Patent ductus arteriosus | 0 (0) | 604 (1.3) | / | / |
| **Respiratory anomalies** | **1 (4.8)** | **1,389 (3.0)** | **1.6 [0.3 - 11.5]** | **1.5 [0.2 - 10.4]** |
| Choanal atresia | 1 (4.8) | 188 (0.4) | 11.9 [1.7 - 85.1] | 9.8 [1.4 - 70.9] |
| **Oro-facial clefts** | **10 (47.8)** | **5,816 (12.2)** | **3.9 [2.1 - 7.2]** | **3.6 [1.9 - 6.6]** |
| Cleft palate | 6 (28.7) | 1,947 (4.2) | 6.9 [3.1 - 15.4] | 6.2 [2.8 - 13.9] |
| Cleft lip/Cleft palate with cleft lip | 4 (19.1) | 3,872 (8.3) | 2.3 [0.9 - 6.2] | 2.2 [0.8 - 5.8] |
| **Anomalies of the digestive system** | **7 (33.5)** | **5,927 (12.7)** | **2.7 [1.3 - 5.6]** | **2.5 [1.2 - 5.2]** |
| Atresia of oesophagus with/without tracheo-oesophageal fistula | 0 (0) | 1,064 (2.3) | / | / |
| Congenital absence, atresia and stenosis of duodenum | 0 (0) | 437 (0.9) | / | / |
| Congenital absence, atresia and stenosis of jejunum/ileum/ other specified parts of small intestine | 1 (4.8) | 436 (0.9) | 5.1 [0.7 - 36.6] | 5.1 [0.7 - 36.1] |
| Congenital absence, atresia and stenosis of anus/rectum with/without fistula | 2 (9.6) | 1,160 (2.5) | 3.9 [1.0 - 15.5] | 3.7 [0.9 – 15.0] |
| Hirschprung disease | 0 (0) | 535 (1.1) | / | / |
| Congenital malformations of intestinal fixation | 3 (14.4) | 438 (0.9) | 15.4 [4.9 - 47.9] | 15.5 [4.9 - 48.5] |
| Atresia of bile ducts | 0 (0) | 263 (0.6) | / | / |
| Annular pancreas | 0 (0) | 23 (0.05) | / | / |
| Congenital diaphragmatic hernia | 1 (4.8) | 939 (2.0) | 2.4 [0.3 - 17.0] | 2.24 [0.3 - 15.9] |
| **Abdominal wall defects** | **1 (4.8)** | **1,256 (2.7)** | **1.8 [0.2 - 12.7]** | **1.8 [0.3 - 12.6]** |
| Gastroschisis | 0 (0) | 640 (1.4) | / | / |
| Exomphalos | 1 (4.8) | 686 (1.5) | 3.3 [0.5 - 23.2] | 2.9 [0.4 - 20.9] |
| **Congenital anomalies of kidney and urinary tract** | **11 (52.6)** | **14,455 (30.9)** | **1.7 [0.9 - 3.1]** | **1.7 [0.9 - 3.1]** |
| Unilateral renal agenesis | 1 (4.8) | 1,629 (3.5) | 1.4 [0.2 - 9.8] | 1.3 [0.2 - 9.5] |
| Bilateral renal agenesis/Potter syndrome | 0 (0) | 89 (0.2) | / | / |
| Renal dysplasia | 2 (9.6) | 1083 (2.3) | 4.1 [1.0 - 16.6] | 3.5 [0.9 - 13.9] |
| Congenital hydronephrosis/Atresia and stenosis of ureter/Other obstructive defects of renal pelvis and ureter | 3 (14.4) | 6,819 (14.6) | 1.0 [0.3 - 3.1] | 1.0 [0.3 - 3.1] |
| Lobulated, fused and horseshoe kidney/Ectopic kidney | 0 (0) | 922 (2.0) | / | / |
| Epispadias/ Exstrophy of urinary bladder | 0 (0) | 151 (0.3) | / | / |
| Congenital posterior urethral valves | 0 (0) | 495 (1.1) | / | / |
| Prune belly syndrome | 0 (0) | 41 (0.09) | / | / |
| **Genital** | **19 (90.9)** | **11,892 (25.4)** | **3.6 [2.3 - 5.7]** | **3.8 [2.4 - 5.9]** |
| Hypospadias | 19 (90.9) | 10,867 (23.2) | 3.9 [2.5 - 6.2] | 4.1 [2.6 - 6.5] |
| Indeterminate sex and pseudohermaphroditism | 0 (0) | 432 (0.9) | / | / |
| **Limb anomalies** | **12 (57.4)** | **13377 (28.6)** | **2.0 [1.1 - 3.6]** | **1.9 [1.1 - 3.4]** |
| Reduction defects of upper/lower/unspecified limb | 0 (0) | 1,002 (2.1) | / | / |
| Talipes equinovarus | 5 (23.9) | 3,915 (8.4) | 2.9 [1.2 – 6.9] | 2.6 [1.1 - 6.2] |
| Congenital dislocation of hip,<br>unilateral/bilateral/unspecified | 1 (4.8) | 3,339 (7.1) | 0.7 [0.1 - 4.8] | 0.7 [0.1 - 4.7] |
| Polydactyly | 6 (28.7) | 4,400 (9.4) | 3.1 [1.4 - 6.8] | 3.0 [1.3 - 6.7] |
| Syndactyly | 0 (0) | 649 (1.4) | / | / |
| ***Other anomalies*** | **24 (114.8)** | **8,313 (17.8)** | **6.5 [4.4 - 9.8]** | **5.5 [3.6 - 8.2]** |
| Craniosynostosis | 11 (52.6) | 1,726 (3.7) | 14.4 [7.9 – 26.0] | 13.2 [7.2 – 24.0] |
| Situs inversus | 0 (0) | 360 (0.8) | / | / |
| Septo-optic dysplasia | 0 (0) | 80 (0.2) | / | / |
| Laterality anomalies | 0 (0) | 622 (1.3) | / | / |

## Discussion

This study reports figures of MCMs prevalence and their associations with prenatal valproate exposure obtained based on data of the EPI-MERES register, a large dataset covering 10.5 million births, i.e. almost all births in France between 2010 and 2023. Information available in the dataset allowed to identify a total of 213,153 live births cases with MCMs overall, and to assess 72 specific MCMs of the EUROCAT list.

Among live births, MCMs prevalence rates estimated in EPI-MERES were generally consistent with those reported in EUROCAT. Overall, among the 10.5 million children from EPI-MERES born between 2010 and 2023 we found a prevalence of any MCM of 203.0 per 10,000 births, very close to the prevalence of 204.8 per 10,000 births reported in EUROCAT between 2010 and 2022. A similar figure (202.5 cases per 10,000 births) was obtained in EPI-MERES when considering a time period strictly comparable to the period of EUROCAT data availability, i.e. 2010-2022. Such consistency underlies the accuracy of the MCMs case-identification algorithms developed in EPI-MERES using information on both hospital diagnoses and medical procedures codes. Medical procedure codes, in particular codes of malformation correction surgery (e.g. surgery for urinary malformation, cleft palate and/or cleft lip, congenital heart defects…), were considered as identification criteria for as many as 22 of the 72 MCMs. Based on these criteria, 47% of all MCM cases were identified based on the presence of a medical procedure code, highlighting the importance of inputting information on medical procedures for MCM identification based on data from healthcare databases.

Based on EPI-MERES data, we found an increase in MCMs prevalence reaching +30.0 cases per 10,000 births, i.e. +16% over the 14-year period between 2010-12 and 2022-2023. This increase was primarily driven by the increasing prevalence of congenital heart defects (+16.6 per 10,000 births), especially atrial septal defect (+12.4 per 10,000 births). This finding is consistent with the general increasing prevalence of mild congenital heart defects resulting from the greater and improving use in echocardiographic techniques over time ^21^, and thus likely reflects an improvement in MCMs identification over time rather than a true increase in their frequency.

Regarding the prevalence estimates for each of the 13 MCMs groups and 72 specific MCMs, some discrepancies were observed between EPI-MERES and EUROCAT. Reassuringly, most of these discrepancies can be explained by several differences in the criteria used for MCMs grouping or definition, such differences being inevitable considering the different nature of the information available in both sources. First, congenital malformation syndromes (ICD-10 codes Q86 and Q87) were included in the group “Other anomalies” in EPI-MERES but in the group “Chromosomal and genetic anomalies” in EUROCAT, resulting in a twice lower prevalence of “Chromosomal and genetic anomalies” in EPI-MERES compared to EUROCAT. Considering ICD-10 codes Q86 and Q87 in the “Chromosomal and genetic anomalies” category would have increased the prevalence of this group from 12.9 to 19.6 cases per 10,000 births. Second, while in EPI-MERES malformations coded as “unspecified” or “other” were not accounted for in the various MCMs groups because such types of codes in healthcare databases may refer to minor congenital anomalies, they are recorded as MCMs in EUROCAT. Third, for 9 MCMs identified using four-digit ICD-10 codes in EUROCAT, identification criteria in EPI-MERES had to be adapted since only three-digit ICD-10 codes are available in the SNDS. For instance, unlike in EUROCAT the definition of atrial septal defect does not exclude persistent foramen ovale (ICD-10 code Q2111) in EPI-MERES, resulting in a 58% higher prevalence estimate for this MCM in EPI-MERES compared to EUROCAT. Fourth, as generally done in registries, in EPI-MERES most MCMs were identified based on diagnoses up to 1 year of age. However, for 3 MCMs for which late diagnosis is common, diagnoses up to age 2 years were considered, increasing the number of detected cases. As an example, 63.3% of hypospadias cases were identified between the age of 1 and 2 years when considering specific hypospadias surgery during the second year of life. Lastly, in addition to differences in MCMs definition criteria, discrepancies between EPI-MERES and EUROCAT may have arisen from differences in terms of covered populations. For instance, the 36% lower prevalence of Down syndrome found in EPI-MERES as compared to EUROCAT likely results from the particularly high frequency of Down syndrome-specific elective pregnancy termination in France, reaching 68% (versus 54% on average in Europe) ^22^.

In our proof-of-concept study on valproate we found that, consistently with results of a recent Cochrane systematic review ^5^, prenatal exposure to valproate was associated with an overall 2.8-fold increase in the risk of occurrence of any MCM. Similarly, as previously reported marked associations were observed with various MCMs organ groups (risk increased by 3.2 for congenital heart defects, by 13.8 for anomalies of the ear, face and neck, by 3.6 for oro-facial clefts, by 1.9 for limb anomalies) and with spina bifida (17.9-fold increased risk). Furthermore, significant associations not reported in the Cochrane review were detected in our study, including associations with specific cardiac anomalies (ventricular septal defect: aOR 3.3; atrial septal defect: aOR 4.4; tetralogy of Fallot: aOR 4.2), with anomalies of the digestive system (aOR 2.5) and with craniosynostosis (aOR 13.2). An association with hypospadias (aOR 4.1) was also identified, corroborating findings from other studies ^17,23,24^. Altogether these findings show not only the interest of the EPI-MERES dataset to accurately measure MCMs, but also its ability, thanks to its particularly large size, to be used to detect differential risks in such rare outcomes as specific MCMs. Actually, while the Cochrane systematic review including all available studies on valproate teratogenicity was based on 4,500 valproate-exposed births overall, they were as many as 2,091 in EPI-MERES alone.

Among births resulting from terminated pregnancies and stillbirths, the MCMs prevalence estimates obtained in EPI-MERES and their comparison with EUROCAT suggest that unlike for live births, information available in EPI-MERES does not allow the correct identification of MCMs among these cases. To address this limitation, in a sensitivity analysis of the study of associations with prenatal valproate exposure we considered as a proxy of any MCM a composite outcome combining live births with any MCM, births resulting from terminated pregnancies and stillbirths. Using this composite outcome, the prevalence of any MCM was 858.0 cases per 10,000 births, close to the rate of 980 per 10,000 reported in the aforementioned Cochrane review ^5^, and the association with prenatal valproate exposure remained similar to that observed in the main analysis. According to the French Biomedicine Agency, the majority (63.3%) of pregnancy interruption after 22 weeks of amenorrhea are related to the identification of a MCM ^25^.

In conclusion, by finding consistent figures of MCMs prevalence and expected associations with prenatal valproate exposure, this study supports the validity of MCMs identification based on data of the EPI-MERES register. Thanks to its large size and to the availability of comprehensive information on prenatal drug exposures during pregnancy, the EPI-MERES register provides a highly powerful, reactive and operational tool complementing MCMs registries for improving knowledge on drug teratogenicity.

## Data Availability

Access to SNDS data is governed by French law and subject to authorization by the French Data Protection Authority (Commission Nationale de l'Informatique et des Libertes, CNIL). The data are not publicly available but may be accessed by researchers meeting the legal and ethical criteria for access to French health administrative data, through application to the CNAM and CNIL.
EPI-PHARE has direct access to the SNDS from the permanent regulatory access of its constitutive bodies, the French National Agency for the Safety of Medicines (ANSM) and Health Products and the French National Health Insurance (Cnam). Permanent access is given according to French Decree No. 2016-1871 of December 26, 2016, relating to the processing of personal data called the "National Health Data System" and French law articles Art. R. 1461-13 and 1461-14. All requests in the database were made by duly authorized persons.

## Statements and Declarations

### Data availability statement

EPI-PHARE has direct access to the SNDS from the permanent regulatory access of its constitutive bodies, the French National Agency for the Safety of Medicines (ANSM) and Health Products and the French National Health Insurance (Cnam). Permanent access is given according to French Decree No. 2016-1871 of December 26, 2016, relating to the processing of personal data called the “National Health Data System” and French law articles Art. R. 1461-13 and 1461-14. All requests in the database were made by duly authorized persons.

### Ethics approval statement

This is an observational study. EPI-PHARE has permanent access to the SNDS database in application of the French Data Protection Authority (CNIL) according to decision CNIL-2016-316. This work was declared, prior to its initiation, on the EPI-PHARE registry of SNDS-based studies (Reference T-2022-03-454).

### Patient Consent statement

No informed consent was required to perform this study. The data used in this study are pseudonymized.

### Authors Contributions

All authors contributed to the study conception and design. Rosemary Dray-Spira designed and directed the project. Data collection was performed by Jerome Drouin, Tom Duchemin and Lise Marty. Material preparation and analysis were performed by Tom Duchemin and Lise Marty. The first draft of the manuscript was written by Tom Duchemin and all authors commented on previous versions of the manuscript. All authors read and approved the final manuscript.

## References

1. World Health Organization. Birth Defects Surveillance: A Manual for Programme Managers, 2nd Edition (No. Cence: CC BY-NC-SA 3.0 IGO). https://www.who.int/publications/i/item/9789240015395 (2020).

2. Boyd, P. A. et al. Paper 1: The EUROCAT network--organization and processes. Birt. Defects Res. A. Clin. Mol. Teratol. 91 **Suppl 1**, S2–15 (2011).

3. Perin, J. et al. Systematic estimates of the global, regional and national under-5 mortality burden attributable to birth defects in 2000-2019: a summary of findings from the 2020 WHO estimates. BMJ Open 13, e067033 (2023).

4. Jentink, J. et al. Valproic acid monotherapy in pregnancy and major congenital malformations. N. Engl. J. Med. 362, 2185–2193 (2010).

5. Bromley, R. et al. Monotherapy treatment of epilepsy in pregnancy: congenital malformation outcomes in the child. Cochrane Database Syst. Rev. 8, CD010224 (2023).

6. Broe, A., Damkier, P., Pottegård, A., Hallas, J. & Bliddal, M. Congenital Malformations in Denmark: Considerations for the Use of Danish Health Care Registries. Clin. Epidemiol. 12, 1371–1380 (2020).

7. Metcalfe, A., Sibbald, B., Lowry, R. B., Tough, S. & Bernier, F. P. Validation of congenital anomaly coding in Canada’s administrative databases compared with a congenital anomaly registry. Birt. Defects Res. A. Clin. Mol. Teratol. 100, 59–66 (2014).

8. Salemi, J. L. et al. The Accuracy of Hospital Discharge Diagnosis Codes for Major Birth Defects: Evaluation of a Statewide Registry With Passive Case Ascertainment. J. Public Health Manag. Pract. JPHMP 22, E9–E19 (2016).

9. Segovia Chacón, S., Karlsson, P. & Cesta, C. E. Detection of major congenital malformations depends on length of follow-up in Swedish National Health Register Data: Implications for pharmacoepidemiological research on medication safety in pregnancy. Paediatr. Perinat. Epidemiol. 38, 521–531 (2024).

10. Nava de Escalante, Y., et al. Validation of case definition algorithms for the ascertainment of congenital anomalies. Birth Defects Res. 115, 302–317 (2023).

11. Tuppin, P. et al. Value of a national administrative database to guide public decisions: From the système national d’information interrégimes de l’Assurance Maladie (SNIIRAM) to the système national des données de santé (SNDS) in France. Rev. Epidemiol. Sante Publique 65 **Suppl 4**, S149–S167 (2017).

12. Miranda, S. et al. Database profile: EPI-MERES, a nationwide mother-child register built from the French National Health Data System (SNDS) for pregnancy and paediatric pharmacoepidemiological research. (2023).

13. Meyer, A. et al. Benefits and Risks Associated With Continuation of Anti-Tumor Necrosis Factor After 24 Weeks of Pregnancy in Women With Inflammatory Bowel Disease : A Nationwide Emulation Trial. Ann. Intern. Med. 175, 1374–1382 (2022).

14. Rios, P. et al. Medically Assisted Reproduction and Risk of Cancer Among Offspring. JAMA Netw. Open 7, e249429 (2024).

15. Tran, A. et al. First-trimester exposure to macrolides and risk of major congenital malformations compared with amoxicillin: A French nationwide cohort study. PLoS Med. 22, e1004576 (2025).

16. Shahriari, P. et al. Trends in Prenatal Exposure to Antiseizure Medications Over the Past Decade: A Nationwide Study. Neurology 105, e213933 (2025).

17. Blotière, P.-O. et al. Risks of 23 specific malformations associated with prenatal exposure to 10 antiepileptic drugs. Neurology 93, e167–e180 (2019).

18. Bergman, J. E. H. et al. Updated EUROCAT guidelines for classification of cases with congenital anomalies. Birth Defects Res. 116, e2314 (2024).

19. Garne, E. et al. Prenatal diagnosis of severe structural congenital malformations in Europe. Ultrasound Obstet. Gynecol. Off. J. Int. Soc. Ultrasound Obstet. Gynecol. 25, 6–11 (2005).

20. Shahriari, P. et al. Trends in prenatal exposure to anti-seizure medications over the past decade: a nationwide study. Neurology (In Press).

21. Liu, Y. et al. Global birth prevalence of congenital heart defects 1970-2017: updated systematic review and meta-analysis of 260 studies. Int. J. Epidemiol. 48, 455–463 (2019).

22. de Graaf, G., Buckley, F. & Skotko, B. G. Estimation of the number of people with Down syndrome in Europe. Eur. J. Hum. Genet. EJHG 29, 402–410 (2021).

23. Veroniki, A. A. et al. Comparative safety of anti-epileptic drugs during pregnancy: a systematic review and network meta-analysis of congenital malformations and prenatal outcomes. BMC Med. 15, 95 (2017).

24. Rodríguez-Pinilla, E. et al. Risk of hypospadias in newborn infants exposed to valproic acid during the first trimester of pregnancy: a case-control study in Spain. Drug Saf. 31, 537–543 (2008).

25. Agence de la Biomédecine. Rapport d’activité Des Centre Pluridisciplinaires de Diagnostic Prénatal. (2022).

