## Supplemental Material for "Identification of Major Congenital Malformations based on Healthcare Databases in France: a Proof-of-Concept Study using the EPI-MERES Nationwide Mother-Child Register"

***Table S1 – Differences in ICD-10 codes used for MCMs identification in EPI-MERES and in EUROCAT***

| Definition in EUROCAT | | Definition in EPI-MERES | | Reason for the difference between EPI-MERES and EUROCAT |
|---|---|---|---|---|
| **MCM** | **ICD-10 codes** | **MCM** | **ICD-10 codes** | |
| Atrial septal defect, excluding persistent foramen ovale | Q211, excluding Q2111 | Atrial septal defect, including persistent foramen ovale | Q211 | 4-digit code Q2111 (persistent foramen ovale) not available in EPI-MERES |
| Agenesis of corpus callosum | Q0400 | Malformations of corpus callosum | Q040 | 4-digit code Q0400 not available in EPI-MERES |
| Arhinencephaly/Holoprencephaly | Q041, Q042, Q8703 | Arhinencephaly/Holoprencephaly | Q041, Q042 | 4-digit code Q8703 (Cyclops syndrome) not available in EPI-MERES |
| Tetralogy and Pentalogy of Fallot | Q213, Q2182 | Tetralogy of Fallot | Q213 | 4-digit code Q2182 (pentalogy of fallot) not available in EPI-MERES |
| Multicystic renal dysplasia | Q6140, Q6141 | Renal dysplasia | Q614 | 4-digit codes Q6140 and Q6141 not available in EPI-MERES |
| Posterior urethral valves | Q6420 | Congenital posterior urethral valves | Q642 | 4-digit code Q6420 not available in EPI-MERES |
| Vascular disruption anomalies | Q0435, Q411, Q412, Q418, Q710, Q712, Q7180, Q720, Q722, Q7280, Q730, Q793, Q7980, Q7982, Q8706 | Vascular disruption anomalies | Q411, Q412, Q418, Q710, Q712, Q720, Q722, Q730, Q793 | 4-digit codes Q0435, Q7180, Q7280, Q7980, Q7982 and Q8706 not available in EPI-MERES |
| Laterality anomalies | Q206, Q240, Q3381, Q890, Q893 | Laterality anomalies | Q206, Q240, Q890, Q893 | 4-digit code Q3381 not available in EPI-MERES |

| | | | | |
|---|---|---|---|---|
| Skeletal dysplasia | Q7402, Q77, Q780-Q788 | Skeletal dysplasia | Q77, Q780-Q788 | 4-digit code Q7402 not available in EPI-MERES |

*Table S2 –List of diagnosis (ICD-10-coded) and medical procedures (CCAM-coded) codes used for identification of Major Congenital Malformations in EPI-MERES*

| | **ICD-10 diagnosis codes** | **CCAM codes** |
|---|---|---|
| **Anomalies of the nervous system** | | |
| Anencephaly and similar malformations | Q00 | |
| Encephalocele | Q01<br>exclude if associated with Anencephaly (Q00) | |
| Spina Bifida | Q05, exclude if associated with Anencephaly (Q00) or Encephalocele (Q01) | |
| Congenital hydrocephalus | Q03<br>exclude if associated with Neural Tube defect group (Q00, Q01, Q05) | |
| Microcephaly | Within 2 years after birth : Q02 + at least 1 MRI within 2 years or death in the six months after birth<br>Exclude if associated with Neural Tube defect group (Q00, Q01, Q05) | ACQJ002, AAQN004, AAQN900, AAQN901, ACQJ001, ACQN004, ACQN001 |
| Arhinencephaly/Holoprosencephaly | Q041, Q042<br>exclude if associated with Neural Tube defect group (Q00, Q01, Q05) | |
| Congenital malformations of corpus callosum | Q040<br>exclude if associated with Neural Tube defect group (Q00, Q01, Q05) | |
| Other specific anomalies of the nervous system | Q045, Q060, Q061, Q062, Q064, Q070 | |

| | | |
|---|---|---|
| Other unspecified anomalies of the nervous system | Q043, Q048, Q049, Q063, Q068, Q069, Q079 | |
| **Anomalies of the eyes** | | |
| Cystic eyeball/Other anophthalmos/Microphthalmos | Q110, Q111, Q112 | |
| Congenital cataract | Q120 + specific medical procedures within 1 year or death within 6 months | BFGA002, BFGA008, BFPA002, BGFA001, BGFA008 |
| Congenital glaucoma | Q150 + specific medical procedures within 1 year or death within 6 months | BEFA008, BEPA003, BGFA014, BHQP002 |
| Other specific anomalies of the eyes | Q100, Q104, Q107, Q113, Q121, Q122, Q123, Q124, Q130, Q131, Q133, Q140, Q141, Q142, Q143 | |
| Other unspecified anomalies of the eyes | Q106, Q128, Q129, Q132, Q134, Q138, Q139, Q148, Q149, Q158, Q159 | |
| **Anomalies of the ear, face and neck** | | |
| Congenital absence of (ear) auricle/Congenital absence atresia and structure of auditory canal (external) | Q160, Q161 | |
| Other specific anomalies of the ear, face and neck | Q162, Q163, Q165, Q183 | |
| Other unspecified anomalies of the ear, face and neck | Q164, Q169, Q178 | |
| **Congenital heart defects** | | |
| Common arterial trunk | Q200 | |
| Double outlet right ventricle | Q201 | |
| Double outlet left ventricle | Q202 | |

| | | |
|---|---|---|
| Discordant ventriculoarterial connection | Q203 | |
| Discordant atrioventricular connection | Q205<br>+ surgical repair within 1 year OR death within 6 months | DZMA001, DZMA002, DZMA003, DZMA004, DZMA006, DZMA010 |
| Double inlet ventricle | Q204<br>exclude if associated with Hypoplastic left heart syndrome (Q234) or Hypoplastic right heart syndrome (Q226) | |
| Ventricular septal defect | Q210 or surgical repair within 1 year | DASA001, DASA004, DASA006, DASA007, DASA009, DASA010, DASA011, DASA012, DASA014, DASF003, DFGA002, DFMA011, DFMA012, DZMA001, DZMA002, DZMA003, DZMA004, DZMA010, DFGA004, DFMA012 |
| Atrial septal defect, incl. persistent foramen ovale | Q211<br>+ at least one echography within 1 year OR death within 6 months | DZQJ001, DZQJ006, DZQJ008, DZQJ009, DZQJ010, DZQJ011, DZQM005, DZQM006 |
| Atrioventricular septal defect | Q212 | |
| Tetralogy of Fallot | Q213 | |
| Congenital tricuspid stenosis | Q224 | |
| Ebstein anomaly | Q225 | |
| Congenital pulmonary valve stenosis | Q221 | |
| Pulmonary valve atresia | Q220 | |
| Congenital stenosis of aortic valve | Q230 | |
| Congenital mitral stenosis | Q232<br>+ surgical repair within 1 year OR death within 6 months | DBMA002, DBMA003 |
| Hypoplastic left heart syndrome | Q234 | |
| Hypoplastic right heart syndrome | Q226 | |
| Coarctation of aorta | Q251<br>+ medical procedure within 1 year OR death within 6 months | DGAA002, DGAA003, DGAA004, DGAA005, DGAA006, DGAF004, DGAF006, DGFA020, DGKA017, DGKA021, DGKA022, DGKA024, EQLF005 |
| Atresia of aorta | Q252 | |

| | | |
|---|---|---|
| Total anomalous pulmonary venous connection | Q262 | |
| Patent ductus arteriosus | Q250<br>+ surgical closure within 1 year OR Q250 still present after 6 months OR death within 6 months<br>exclude if associated with transposition of great arteries (Q203), hypoplastic left heart (Q234) and coarctation of aorta (Q251) | DASF001 |
| Other specific congenital heart defects | Q206, Q214, Q222, Q231, Q233, Q240, Q241, Q242, Q243, Q244, Q245, Q253, Q255, Q256, Q260, Q263, Q265, Q266 | |
| Other unspecified congenital heart defects | Q208, Q209, Q218, Q219, Q223, Q228, Q229, Q238, Q239, Q248, Q249, Q257, Q258, Q259, Q264, Q268, Q269 | |
| **Respiratory anomalies** | | |
| Choanal atresia | Q300<br>+ surgical repair within 1 year or death within 6 months | GCCD001, GCMA001, GCME001, GCME002, GCME003, GCME004 |
| Other specific respiratory anomalies | Q323, Q332, Q333, Q334, Q335, Q336, Q340, Q341 | |
| Other unspecified respiratory anomalies | Q321, Q324, Q338, Q339, Q348, Q349 | |
| **Oro-facial clefts** | | |
| Cleft palate | Q35<br>exclude Q357 and exclude if associated with holoprosencephaly subgroup, or cleft lip subgroups | |
| Cleft lip/Cleft palate with cleft lip | Q36, Q37<br>exclude if associated with holoprosencephaly subgroup | |
| **Anomalies of the digestive system** | | |

| | | |
|---|---|---|
| Atresia of oesophagus with/without tracheo-oesophageal fistula | Q390, Q391 | |
| Congenital absence, atresia and stenosis of duodenum | Q410<br>exclude if associated with annular pancreas subgroup | |
| Congenital absence, atresia and stenosis of jejunum/ileum/ other specified parts of small intestine | Q411, Q418 | |
| Congenital absence, atresia and stenosis of anus/rectum with/without fistula | Q420, Q421, Q422, Q423<br>+ surgical repair within 1 year or death within 6 months | HHCA002, HHCC007, HJAD001, HJEA001, HJEA002, HJEA003, HJEA004, HJMA001, HKEA001, HKMA006 |
| Hirschprung disease | Q431 + surgical repair within 1 year or death within 6 months | HHCA002, HHCC007, HJFA016, HJFC001, HJFD003 |
| Congenital malformations of intestinal fixation | Q433 | |
| Atresia of bile ducts | Q442 + surgical repair within 1 year or death within 6 months | HLCA001 |
| Annular pancreas | Q451 | |
| Congenital diaphragmatic hernia | Q790 | |
| Other specific anomalies of the digestive system | Q384, Q387, Q392, Q393, Q394, Q395, Q396, Q434, Q435, Q436, Q437, Q440, Q443, Q446, Q450, Q452 | |
| Other unspecified anomalies of the digestive system | Q380, Q383, Q386, Q388, Q398, Q399, Q402, Q403, Q408, Q409, Q419, Q428, Q429, Q439, Q441, Q445, Q447, Q453, Q459 | |
| **Abdominal wall defects** | | |
| Gastroschisis | Q793 | |
| Exomphalos | Q792 | |
| Other unspecified abdominal anomalies | Q795 | |

| | | |
|---|---|---|
| **Congenital anomalies of kidney and urinary tract** | | |
| Unilateral renal agenesis | Q600 | |
| Bilateral renal agenesis/Potter syndrome | Q601, Q606 | |
| Renal dysplasia | Q614 | |
| Congenital hydronephrosis/Atresia and stenosis of ureter/Other obstructive defects of renal pelvis and ureter | Q620, Q621, Q623<br>+ at least 2 echographies within 1 year or death within 6 months<br>exclude if associated with Q627 | JAQJ001, JAQM001, JAQM003, JAQM004 |
| Lobulated, fused and horseshoe kidney/Ectopic kidney | Q631, Q632 | |
| Epispadias/ Exstrophy of urinary bladder | Within 2 years after birth: Q640<br>+ surgical repair within 1 or 2 year(s) or death within 6 months<br>OR<br>Q641<br>+ surgical repair within 1 or death within 6 months | JHAA001, JHAA002<br><br>JDFA010, JDFA012, JDFA013, JDSA001, JDSA003, JDSA004, JDSA007, JDSA009 |
| Congenital posterior urethral valves | Q642<br>+ surgical repair within 1 year or death within 6 months | JEFE005, JEPH001 |
| Prune belly syndrome | Q794 | |
| Other specific kidney and urinary tract anomalies | Q603, Q604, Q611, Q612, Q615, Q618, Q622, Q624, Q625, Q626, Q630, Q644, Q645, Q646 | |
| Other unspecified kidney and urinary tract anomalies | Q602, Q605, Q613, Q619, Q628, Q638, Q639, Q643, Q647, Q648, Q649 | |
| **Genital** | | |
| Hypospadias | Within 2 years after birth: Q54 excluding Q544<br>+ surgical repair within 1 or 2 years | JEMA006, JEMA011, JEMA014, JEMA019, JEMA020, JEMA021 |

| | | |
|---|---|---|
| Indeterminate sex and pseudohermaphroditism | Q56 | |
| Other specific genital anomalies | Q500, Q504, Q510, Q511, Q513, Q514, Q515, Q516, Q517, Q520, Q521, Q522, Q526, Q550, Q551, Q553, Q555 | |
| Other unspecified genital anomalies | Q503, Q506, Q512, Q518, Q519, Q524, Q528, Q529, Q554, Q556, Q558, Q559 | |
| **Limb anomalies** | | |
| Reduction defects of upper/lower/unspecified limb | Q71, Q72, Q73 | |
| Talipes equinovarus | Q660<br>+ specific medical procedures within 1 year OR death within 6 months | NHRP003, NJAB001, PCPB002 |
| Congenital dislocation of hip, unilateral/bilateral/unspecified | Q650, Q651, Q652<br>+ surgical repair within 1 year OR at least 2 diagnostic tests within 1 year OR death within 6 months | Surgery: NEEP003, NEEA004, ZEMP002, NEQP001, NEQH001, NEEP006, NEQP002, NZMP012, ZEMP010<br><br>Diagnostic test: NEQM001, NEQH002, NAQK071, NEQC001 |
| Polydactyly | Q69<br>+ surgical repair within 1 year OR death within 6 months | MZFA008, MZFA012, MZFA014, MZFA015, NZFA011, NZFA012 |
| Syndactyly | Q70<br>+ surgical repair within 1 year OR death within 6 months | MJPA014, MZPA002, QDPA001 |
| Other specific limb anomalies | Q741, Q743 | |
| Other unspecified limb anomalies | Q688, Q742, Q748, Q749 | |
| **Other anomalies** | | |
| Craniosynostosis | Q750<br>+ surgical repair within 1 year or death within 6 months | LAEA002, LAEA004, LAEA006, LAEA009, LAFA900, LAMA006, LANC001, LAPA005, LAPA006, LAPA008, LAPA016, LARA001, LARA002, LARA003, LARA004 |

| | |
|---|---|
| Situs inversus<br>Septo-optic dysplasia<br>Laterality anomalies | Q893<br>Q044<br>Q206, Q240, Q890, Q893 |
| Other specific anomalies | D215, Q271, Q273, Q274, Q280, Q282, Q301, Q302, Q303, Q310, Q311, Q312, Q313, Q755, Q761, Q762, Q763, Q830, Q831, Q832, Q840, Q841, Q843, Q844, Q892, Q870, Q871, Q872, Q873, Q874, Q894, Q897, Q86, P350 |
| Other unspecified anomalies | Q272, Q278, Q279, Q281, Q283, Q288, Q289, Q308, Q309, Q318, Q319, Q758, Q759, Q768, Q769, Q791, Q799, Q829, Q838, Q839, Q842, Q848, Q849, Q858, Q859, Q875, Q878, Q898 |
| **Chromosomal**<br>Skeletal dysplasia<br>Down syndrome<br>Trisomy 13/Patau syndrome<br>Trisomy 18/Edwards syndrome<br>Turner syndrome<br>Triploidy and polyploidy | <br>Q77, Q780, Q781, Q782, Q783, Q784, Q758, Q786, Q788<br>Q90<br>Q914, Q915, Q916, Q917<br>Q910, Q911, Q912, Q913<br>Q96<br>Q927 |
| Other specific chromosomal anomalies | D821, Q751, Q754, Q796, Q800, Q801, Q802, Q803, Q804, Q810, Q811, Q812, Q820, Q821, Q822, Q823, Q824, Q850, Q851, Q920, Q921, Q922, Q923, Q924, Q925, Q926, Q930, Q931, Q932, Q933, Q934, Q936, Q970, Q971, Q972, Q973, Q980, Q981, Q982, Q985, Q986, Q987, Q990, Q991, Q992 |
| Other unspecified chromosomal anomalies | Q789, Q808, Q809, Q818, Q819, Q928, Q929, Q935, Q937, Q938, Q939, Q978, Q979, Q983, Q984, Q988, Q989, Q998, Q999 |

**Table S3 – Teratogenic drugs**

| Anatomical Therapeutic Chemical (ATC) Classification System | Therapeutic class | Drug |
|---|---|---|
| A02BB01 | Prostaglandins | Misoprostol |
| B01AA02<br>B01AA03<br>B01AA07<br>B01AA12 | Vitamin K antagonists | Phenindione<br>Warfarin<br>Acenocoumarol<br>Fluindione |
| D05AX05 | Antipsoriatics for topical use | Tazarotene |
| D05BB02 | Antipsoriatics for systematic use | Acitretin |
| D10BA01 | Retinoids for treatment of acne | Isotretinoin |
| D11AH04 | Agents for dermatitis, oral | Alitretinoin |
| G03XA01<br>G03XC01 | Sex hormones and modulators of the genital system | Danazol<br>Raloxifene |
| G04CB01<br>G04CB02 | Drugs used in benign prostatic hypertrophy | Finasteride<br>Dutasteride |
| J05AP01 | Direct acting antivirals | Ribavirin |
| L01A | Alkylating agents | All alkylating agents |
| L01B (except L01BB02) | Antimetabolites (except Mercaptopurine) | All antimetabolites (except Mercaptopurine) |
| L01C | Plant alkaloids and other natural products | All plant alkaloids and other natural products |
| L01D | Cyctotoxic antibiotics and related substances | All cyctotoxic antibiotics and related substances |
| L01X | Other antineoplastic agents | All other antineoplastic agents |
| L02AB01<br>L02AE03 | Hormones and related agents | Megestrol<br>Goserelin |
| L02BA01<br>L02BA03<br>L02BB03<br>L02BG03<br>L02BG06 | Hormone antagonists and related agents | Tamoxifen<br>Fulvestrant<br>Bicalutamide<br>Anastrozole<br>Exemestane |
| L04AA06<br>L04AA13<br>L04AA31<br>L04AA40 | Immunosuppressants | Mycophenolic acid<br>Leflunomide<br>Teriflunomide<br>Cladribine |

| | | |
|---|---|---|
| L04AX02 | | Thalidomide |
| L04AX03 | | Methotrexate |
| N03AG01 | Antiepileptics | Valproic acid |
| N03AX11 | | Topiramate |

**Table S4 - Algorithm to identify teratogenic infections or suspected teratogenic infections**

| Infection | | Hospital discharge diagnoses codes (International Classification of Diseases, 10th revision) | Drugs (ATC class) |
|---|---|---|---|
| Toxoplasmosis | Mother | Toxoplasmosis (B58) | Pyrimethamine (P01BD01) AND Sulfadiazine (J01EC02) OR Spiramycine (J01FA02) if at least three dispensing during pregnancy |
| | Child | Congenital toxoplasmosis (P371) | Pyrimethamine (P01BD01) AND Sulfadiazine (J01EC02) up to one year of life |
| Syphilis | Mother | Syphilis complicating pregnancy, childbirth, and the puerperium (O981)<br>Early syphilis (A51)<br>Other and unspecified syphilis (A53)<br>Cardiovascular syphilis (I980)<br>Late syphilis of kidney (N290) | Benzylpenicilline (J01CE01) OR benzathine benzylpenicilline (J01CE08) |
| | Child | Congenital syphilis (A50) | Benzylpenicilline (J01CE01) OR benzathine benzylpenicilline (J01CE08) up to one year of life |
| Rubella | Mother | Rubella (B06)<br>Rubella arthritis (M014)<br>Maternal care for damage to fetus from maternal: cytomegalovirus/rubella (O353) | |
| | Child | Congenital rubella syndrome (P350) | |

| | | |
|---|---|---|
| Cytomegalovirus | Mother | Cytomegaloviral disease (B25)<br>Cytomegaloviral mononucleosis (B271)<br>HIV disease resulting in Cytomegaloviral disease (B202)<br>Cytomegaloviral cholangitis (K8700)<br>Cytomegaloviral colitis (K93820)<br>Cytomegaloviral retinitis (H3200)<br>Maternal care for damage to fetus from maternal: cytomegalovirus/rubella (O353) |
| | Child | Congenital cytomegalovirus infection (P351) |
| Herpes | Mother | Herpes gestationis (O264), <u>only PD/RD</u> |
| | Child | Congenital herpes viral [herpes simplex] infection (P352), <u>only PD/RD</u> |
| Varicella | Mother | Varicella (B01) |
| | Child | Congenital varicella (P358) |
| Lymphocytic choriomeningitis virus | Mother | Lymphocytic choriomeningitis (A872) |
| | Child | / |
| Zika | Mother | Zika virus disease (A925) |
| | Child | Congenital Zika virus disease (P354) |

**Table S5 – Variables of the valproate cohort study**

| Category | Variables |
|---|---|
| | **Exposure (ATC codes)** |
| **Valproate** | N03AG01 - Valproic acid (at the exception of Depakote and Divalcote) |
| **Exclusion criteria (other antiepileptic drugs)** | N02BF01 - GABAPENTINE<br>N02BF02 - PREGABALINE<br>N03AA02 - PHENOBARBITAL<br>N03AA03 - PRIMIDONE<br>N03AB02 - PHENYTOINE<br>N03AD01 - ETHOSUXIMIDE<br>N03AE01 - CLONAZEPAM<br>N03AF01 - CARBAMAZEPINE<br>N03AF02 - OXCARBAZEPINE<br>N03AF03 - RUFINAMIDE<br>N03AF04 - ESLICARBAZEPINE<br>N03AG02 - VALPROMIDE<br>N03AG04 - VIGABATRIN<br>N03AG05 - PROGABIDE<br>N03AG06 - TIAGABINE<br>N03AX09 - LAMOTRIGINE<br>N03AX11 - TOPIRAMATE<br>N03AX12 - GABAPENTINE<br>N03AX14 - LEVETIRACETAM<br>N03AX15 - ZONISAMIDE<br>N03AX17 - STIRIPENTOL<br>N03AX18 - LACOSAMIDE<br>N03AX21 - RETIGABINE<br>N03AX22 - PERAMPANEL<br>N03AX23 - BRIVARACETAM |
| | **Covariates of the propensity score** |

| | |
|---|---|
| **Socio-demographic** | - Mother's age at start of pregnancy: <25 years / 25 - 34 years / 35 - 39 years / >39 years<br>- Year pregancy ended in 3 classes: 2010 - 2011 / 2012 - 2013 / 2014 - 2015<br>- Pregnancy rank among all pregnancies since 2010<br>- Sex of the child<br>- 2015 Deprivation index of the commune of residence at the start of the pregancy and categorized in quintiles<br>- Level of resources as a combination on CMU-c and the level of gross monthly salary calculated from the amount of daily maternity benefit: (1) CMU-C, (2) Salary < median salary and no CMU-c, (3) Salary >= median salary and no CMU-c, (4) No CMU-c and unknown salary<br>- Localized potential access of the commune of residence: (1) >4 consultations/year/inhabitant, (2) 2.5- 4 consultations/year/inhabitant, (3) <=2.5 consultations/inhabitant/year |
| **Comorbidities** | - Psychatric pathologies of the mother<br>- Maternal diabetes : (1) no or uncertain diabetes, (2) gestational diabetes, (3) type 1 or type 2 diabetes<br>- Pre-gestational hypertention<br>- Presence of obesity indicator (hospitalization with an ICD-10 code for obesity or a long-term disease indicator in the previous 5 years) |
| **Lifestyle** | - Presence of tobacco consumption indicators<br>- Presence of opiate consumption indicators<br>- Presence of alcohol consumption indicators. |
| **Drug exposure** | - Folic acid supplementation during the periconceptional period (between 61 days and 91 days after the date of conception for normal pack / between 122 days before and 62 days after for large pack)<br>- Psychotropic drugs (antidepressants, anxiolytics, hypnotics and/or neuroleptics) |

**Table S6 - MCMs prevalence among live births in EPI-MERES and in EUROCAT**

| | Prevalence (live births per 10,000 live- and stillbirths) | | | | | | | |
|---|---|---|---|---|---|---|---|---|
| | EPI-MERES | | | | | | EUROCAT | Difference in prevalence between EPI-MERES and EUROCAT n (%) |
| **MCM** | 2010 - 2012 | 2013 - 2015 | 2016 - 2018 | 2019 - 2021 | 2022-2023 | All | All | All |
| **Any MCM** | **181.2** | **200.4** | **211.8** | **211.4** | **211.2** | **203.0** | **204.8** | **-1.8 (-1%)** |
| **Anomalies of the nervous system** | **8.9** | **10.6** | **11.0** | **12.0** | **13.9** | **11.2** | **12.1** | **-0.9 (-7%)** |
| Anencephaly and similar malformations | 0.04 | 0.13 | 0.15 | 0.14 | 0.17 | 0.12 | 0.20 | -0.1 (-40%) |
| Encephalocele | 0.4 | 0.4 | 0.5 | 0.4 | 0.5 | 0.4 | 0.3 | 0.1 (34%) |
| Spina Bifida | 1.1 | 1.6 | 1.7 | 1.6 | 1.5 | 1.5 | 1.6 | -0.1 (-7%) |
| Congenital hydrocephalus | 2.8 | 2.4 | 2.6 | 1.9 | 2.2 | 2.4 | 2.7 | -0.3 (-11%) |
| Microcephaly | 2.8 | 3.6 | 3.5 | 4.7 | 6.1 | 3.9 | 2.2 | 1.8 (82%) |
| Arhinencephaly/Holoprosencephaly | 0.1 | 0.1 | 0.1 | 0.2 | 0.2 | 0.1 | 0.3 | -0.1 (-50%) |
| Congenital malformations of corpus callosum | 1.6 | 2.3 | 2.4 | 3.2 | 3.3 | 2.6 | 1.2 | 1.4 (111%) |
| **Anomalies of the eyes** | **4.5** | **4.6** | **5.0** | **4.4** | **4.3** | **4.6** | **3.8** | **0.8 (21%)** |
| Cystic eyeball/Other anophthalmos/Microphthalmos | 0.8 | 0.6 | 1.0 | 0.8 | 0.8 | 0.8 | 0.6 | 0.2 (27%) |
| Congenital cataract | 1.4 | 1.4 | 1.6 | 1.3 | 1.6 | 1.4 | 1.3 | 0.1 (6%) |
| Congenital glaucoma | 1.0 | 1.0 | 0.8 | 0.8 | 0.8 | 0.9 | 0.3 | 0.6 (197%) |
| **Anomalies of the ear, face and neck** | **0.6** | **0.7** | **0.8** | **0.8** | **0.6** | **0.7** | **1.5** | **-0.7 (-50%)** |
| Congenital absence of (ear) auricle/Congenital absence atresia and structure of auditory canal (external) | 0.5 | 0.5 | 0.5 | 0.5 | 0.4 | 0.5 | 0.8 | -0.3 (-34%) |
| **Congenital heart defects** | **50.1** | **59.5** | **65.3** | **68.6** | **66.7** | **62.3** | **71.4** | **-9.1 (-13%)** |
| Common arterial trunk | 0.5 | 0.5 | 0.5 | 0.4 | 0.4 | 0.5 | 0.4 | 0.0 (9%) |

| | | | | | | | | |
|---|---|---|---|---|---|---|---|---|
| Double outlet right ventricle | 0.8 | 1.1 | 1.0 | 1.1 | 1.3 | 1.1 | 1.2 | -0.1 (-9%) |
| Double outlet left ventricle | 0.08 | 0.11 | 0.11 | 0.17 | 0.17 | 0.11 | 0.03 | 0.1 (256%) |
| Discordant ventriculoarterial connection | 2.9 | 3.4 | 3.2 | 3.2 | 3.3 | 3.2 | 2.9 | 0.3 (9%) |
| Discordant atrioventricular connection | 0.08 | 0.15 | 0.09 | 0.10 | 0.07 | 0.12 | 0.39 | -0.3 (-70%) |
| Double inlet ventricle | 0.5 | 0.7 | 0.6 | 0.6 | 0.8 | 0.6 | 0.2 | 0.4 (174%) |
| Ventricular septal defect | 26.2 | 29.2 | 31.3 | 30.4 | 28.3 | 29.3 | 36.6 | -7.3 (-20%) |
| Atrial septal defect, incl. persistent foramen ovale | 16.3 | 21.6 | 25.7 | 29.1 | 28.7 | 24.1 | 15.3 | 8.8 (58%) |
| Atrioventricular septal defect | 2.6 | 2.7 | 2.9 | 3.2 | 3.3 | 3.0 | 3.2 | -0.3 (-8%) |
| Tetralogy of Fallot | 3.2 | 3.4 | 3.2 | 3.0 | 3.4 | 3.3 | 3.0 | 0.3 (10%) |
| Congenital tricuspid stenosis | 0.3 | 0.4 | 0.5 | 0.5 | 0.4 | 0.4 | 0.4 | 0.0 (6%) |
| Ebstein anomaly | 0.3 | 0.3 | 0.4 | 0.4 | 0.3 | 0.3 | 0.4 | -0.1 (-14%) |
| Congenital pulmonary valve stenosis | 2.8 | 3.2 | 3.3 | 3.5 | 4.0 | 3.3 | 4.2 | -0.8 (-20%) |
| Pulmonary valve atresia | 0.9 | 1.1 | 1.0 | 1.1 | 1.3 | 1.1 | 0.9 | 0.2 (29%) |
| Congenital stenosis of aortic valve | 1.1 | 1.1 | 1.1 | 0.9 | 0.8 | 1.0 | 1.3 | -0.3 (-23%) |
| Congenital mitral stenosis | 0.1 | 0.3 | 0.2 | 0.2 | 0.3 | 0.2 | 0.3 | -0.1 (-37%) |
| Hypoplastic left heart syndrome | 0.9 | 1.5 | 1.3 | 1.4 | 1.5 | 1.3 | 1.3 | 0.0 (1%) |
| Hypoplastic right heart syndrome | 0.3 | 0.5 | 0.5 | 0.5 | 0.6 | 0.5 | 0.3 | 0.2 (59%) |
| Coarctation of aorta | 3.2 | 3.9 | 3.5 | 3.3 | 3.3 | 3.5 | 3.7 | -0.3 (-7%) |
| Atresia of aorta | 0.3 | 0.5 | 0.5 | 0.4 | 0.6 | 0.5 | 0.4 | 0.1 (14%) |
| Total anomalous pulmonary venous connection | 0.7 | 0.7 | 0.6 | 0.7 | 0.6 | 0.7 | 0.7 | 0.0 (-3%) |
| Patent ductus arteriosus | 1.4 | 1.7 | 1.6 | 1.9 | 1.8 | 1.6 | 2.9 | -1.3 (-44%) |
| **Respiratory anomalies** | **2.9** | **3.0** | **3.5** | **3.3** | **3.7** | **3.4** | **3.3** | **0.1 (3%)** |
| Choanal atresia | 0.3 | 0.5 | 0.4 | 0.4 | 0.5 | 0.4 | 0.9 | -0.4 (-51%) |
| **Oro-facial clefts** | **13.1** | **12.6** | **12.9** | **11.7** | **12.4** | **12.5** | **12.6** | **-0.1 (-1%)** |
| Cleft palate | 4.3 | 4.6 | 4.6 | 4.1 | 4.1 | 4.3 | 7.4 | -3.1 (-42%) |
| Cleft lip/Cleft palate with cleft lip | 8.7 | 8.1 | 8.2 | 7.6 | 8.3 | 8.2 | 5.2 | 3.0 (57%) |
| **Anomalies of the digestive system** | **12.2** | **13.6** | **14.1** | **14.2** | **15.7** | **13.9** | **15.2** | **-1.3 (-8%)** |

| | | | | | | | | |
|---|---|---|---|---|---|---|---|---|
| Atresia of oesophagus with/without tracheo-oesophageal fistula | 2.2 | 2.4 | 2.5 | 2.7 | 3.0 | 2.5 | 2.4 | 0.0 (2%) |
| Congenital absence, atresia and stenosis of duodenum | 1.0 | 1.1 | 1.3 | 1.4 | 1.3 | 1.2 | 1.2 | 0.0 (-1%) |
| Congenital absence, atresia and stenosis of jejunum/ileum/ other specified parts of small intestine | 0.8 | 1.0 | 1.1 | 1.2 | 1.1 | 1.0 | 0.8 | 0.2 (25%) |
| Congenital absence, atresia and stenosis of anus/rectum with/without fistula | 2.4 | 2.7 | 2.9 | 2.6 | 3.5 | 2.8 | 2.7 | 0.1 (3%) |
| Hirschprung disease | 1.3 | 1.2 | 1.3 | 1.1 | 1.3 | 1.2 | 1.4 | -0.2 (-13%) |
| Congenital malformations of intestinal fixation | 0.9 | 1.0 | 0.9 | 1.0 | 1.0 | 1.0 | 0.3 | 0.7 (202%) |
| Atresia of bile ducts | 0.5 | 0.6 | 0.5 | 0.5 | 0.5 | 0.5 | 0.2 | 0.4 (231%) |
| Annular pancreas | 0.06 | 0.03 | 0.07 | 0.09 | 0.07 | 0.07 | 1.11 | -1.0 (-94%) |
| Congenital diaphragmatic hernia | 1.8 | 2.2 | 2.3 | 2.1 | 2.5 | 2.2 | 2.1 | 0.1 (4%) |
| **Abdominal wall defects** | **2.3** | **3.0** | **2.8** | **2.9** | **2.5** | **2.8** | **3.4** | **-0.6 (-18%)** |
| Gastroschisis | 1.3 | 1.3 | 1.4 | 1.4 | 1.2 | 1.4 | 2.1 | -0.7 (-34%) |
| Exomphalos | 1.2 | 1.8 | 1.5 | 1.5 | 1.4 | 1.5 | 1.2 | 0.3 (22%) |
| **Congenital anomalies of kidney and urinary tract** | **29.6** | **31.9** | **34.9** | **33.7** | **31.6** | **32.4** | **30.3** | **2.1 (7%)** |
| Unilateral renal agenesis | 3.3 | 3.5 | 4.1 | 4.3 | 4.7 | 4.0 | 3.4 | 0.6 (16%) |
| Bilateral renal agenesis/Potter syndrome | 0.1 | 0.2 | 0.2 | 0.2 | 0.2 | 0.2 | 0.2 | 0 (-14%) |
| Renal dysplasia | 2.1 | 2.4 | 2.7 | 3.1 | 2.8 | 2.6 | 3.5 | -0.9 (-25%) |
| Congenital hydronephrosis/Atresia and stenosis of ureter/Other obstructive defects of renal pelvis and ureter | 13.7 | 15.2 | 16.6 | 14.8 | 13.0 | 14.8 | 14.5 | 0.3 (2%) |
| Lobulated, fused and horseshoe kidney/Ectopic kidney | 1.7 | 2.1 | 2.7 | 3.3 | 3.2 | 2.6 | 2.7 | -0.1 (-3%) |
| Epispadias/ Exstrophy of urinary bladder | 0.3 | 0.4 | 0.3 | 0.3 | 0.3 | 0.3 | 0.4 | -0.2 (-36%) |
| Congenital posterior urethral valves | 0.9 | 1.1 | 1.4 | 1.1 | 1.3 | 1.2 | 1.0 | 0.1 (14%) |
| Prune belly syndrome | 0.11 | 0.09 | 0.09 | 0.05 | 0.06 | 0.08 | 0.05 | 0.0 (66%) |
| **Genital** | **24.8** | **25.7** | **26.1** | **24.4** | **22.6** | **24.4** | **20.9** | **3.4 (16%)** |

| | | | | | | | | |
|---|---|---|---|---|---|---|---|---|
| Hypospadias | 22.8 | 23.3 | 23.6 | 21.3 | 19.4 | 21.8 | 18.6 | 3.2 (17%) |
| Indeterminate sex and pseudohermaphroditism | 0.9 | 1.0 | 0.9 | 1.0 | 0.7 | 1.0 | 0.3 | 0.6 (181%) |
| **Limb anomalies** | **26.3** | **30.8** | **31.2** | **30.9** | **32.3** | **30.4** | **33.0** | **-2.6 (-8%)** |
| Reduction defects of upper/lower/unspecified limb | 2.1 | 2.4 | 2.3 | 2.4 | 2.1 | 2.3 | 3.2 | -1.0 (-30%) |
| Talipes equinovarus | 7.5 | 9.3 | 9.1 | 9.5 | 9.4 | 9.0 | 9.6 | -0.6 (-6%) |
| Congenital dislocation of hip, unilateral/bilateral/unspecified | 6.7 | 7.3 | 7.1 | 5.8 | 6.0 | 6.7 | 6.0 | 0.6 (11%) |
| Polydactyly | 8.5 | 10.2 | 11.3 | 12.0 | 13.3 | 11.0 | 10.0 | 1.0 (10%) |
| Syndactyly | 1.5 | 1.4 | 1.2 | 0.9 | 1.1 | 1.2 | 3.6 | -2.4 (-67%) |
| **Other anomalies** | | | | | | | | |
| Craniosynostosis | 3.8 | 3.9 | 4.2 | 4.4 | 4.5 | 4.1 | 2.8 | 1.3 (49%) |
| Situs inversus | 0.6 | 0.8 | 0.7 | 1.1 | 0.8 | 0.9 | 0.6 | 0.2 (38%) |
| Septo-optic dysplasia | 0.1 | 0.2 | 0.3 | 0.4 | 0.4 | 0.3 | 0.2 | 0.1 (84%) |
| Laterality anomalies | 1.0 | 1.5 | 1.4 | 1.8 | 1.6 | 1.5 | 1.5 | 0.0 (3%) |
| **Chromosomal** | **10.7** | **12.3** | **13.1** | **13.9** | **15.2** | **12.9** | **25.7** | **-12.8 (-50%)** |
| Skeletal dysplasia | 0.9 | 1.4 | 1.6 | 1.9 | 2.5 | 1.7 | 1.2 | 0.4 (35%) |
| Down syndrome | 5.7 | 6.2 | 6.7 | 6.3 | 7.1 | 6.3 | 9.8 | -3.5 (-36%) |
| Trisomy 13/Patau syndrome | 0.1 | 0.3 | 0.3 | 0.3 | 0.3 | 0.3 | 0.3 | 0.0 (-16%) |
| Trisomy 18/Edwards syndrome | 0.3 | 0.5 | 0.4 | 0.6 | 0.6 | 0.5 | 0.7 | -0.3 (-37%) |
| Turner syndrome | 0.5 | 0.5 | 0.5 | 0.6 | 0.4 | 0.5 | 0.7 | -0.1 (-21%) |
| Triploidy and polyploidy | 0.02 | 0.02 | 0.04 | 0.02 | 0.01 | 0.02 | 0.04 | 0.0 (-38%) |

**Table S7 – Prevalence MCMs per 10,000 live- and stillbirths of the 11 that can be identified among stillbirths and pregnancy interruption after 22 weeks of amenorrhea in EPI-MERES (2010-2023) and EUROCAT (2010-2022)**

| | EPI-MERES | EUROCAT | Difference between EPI-MERES and EUROCAT (%) |
|---|---|---|---|
| Anencephaly and similar malformations | 0.2 | 4.2 | -4.0 (-94%) |
| Encephalocele | 0.5 | 1.2 | -0.7 (-55%) |
| Spina Bifida | 2.8 | 5.0 | -2.2 (-44%) |
| Congenital hydrocephalus | 3.0 | 5.2 | -2.3 (-43%) |
| Discordant ventriculoarterial connection | 3.2 | 3.5 | -0.3 (-8%) |
| Hypoplastic left heart syndrome | 1.7 | 2.8 | -1.1 (-40%) |
| Congenital diaphragmatic hernia | 2.4 | 3.0 | -0.6 (-19%) |
| Gastroschisis | 1.4 | 2.6 | -1.2 (-46%) |
| Exomphalos | 1.6 | 3.9 | -2.3 (-59%) |
| Bilateral renal agenesis/Potter syndrome | 0.5 | 1.3 | -0.8 (-64%) |
| Reduction defects of upper/lower/unspecified limb | 2.4 | 5.1 | -2.7 (-53%) |

*Table S8 – Characteristics of children exposed and unexposed to prenatal valproate monotherapy*

| | Exposed | Unexposed |
|---|---|---|
| n | 2 091 | 4 683 744 |
| **Sociodemographic characteristics** | | |
| **Age** | | |
| *Mean (SD)* | 30.9 (5.8) | 30 (19.4) |
| <25 years old / n (%) | 306 (14.6%) | 752 503 (16.1%) |
| 25 - 34 years old / n (%) | 1 186 (56.7%) | 3 005 683 (64.2%) |
| 35 - 39 years old / n (%) | 462 (22.1%) | 739 289 (15.8%) |
| 40 years old and more / n (%) | 137 (6.6%) | 186 269 (4.0%) |
| **Year of end of pregnancy / n (%)** | | |
| 2010 - 2011 | 938 (44.9%) | 1 571 716 (33.6%) |
| 2012 - 2013 | 662 (31.7%) | 1 567 775 (33.5%) |
| 2014 - 2015 | 491 (23.5%) | 1 544 253 (33.0%) |
| **Sex of the child / n (%)** | | |
| Female | 1 054 (50.4%) | 2 289 122 (48.9%) |
| Male | 1 037 (49.6%) | 2 394 622 (51.1%) |
| **Gravidity since 2010 / n (%)** | | |
| 1st pregnancy | 1 594 (76.2%) | 3 403 012 (72.7%) |
| 2nd pregnancy | 415 (19.9%) | 1 062 343 (22.7%) |
| 3rd pregnancy or more | 82 (3.9%) | 218 389 (4.7%) |
| **Deprivation index / n (%)** | | |
| Quintile 1 (less deprived) | 291 (13.9%) | 862 870 (18.4%) |
| Quintile 2 | 314 (15.0%) | 876 437 (18.7%) |
| Quintile 3 | 385 (18.4%) | 872 332 (18.6%) |
| Quintile 4 | 449 (21.5%) | 865 062 (18.5%) |
| Quintile 5 (more deprived) | 541 (25.9%) | 939 211 (20.1%) |
| Oversea departments and territories | 31 (1.5%) | 78 618 (1.7%) |
| **Income / n (%)** | | |
| Complementary Universal Health Insurance (CMU-c / C2S) | 578 (27.6%) | 706 158 (15.1%) |
| Monthly wage < median wage and no CMU-c/C2S | 524 (25.1%) | 1 112 920 (23.8%) |
| Monthly wage >= median wage and no CMU-c | 301 (14.4%) | 1 224 430 (26.1%) |
| No or unknown wage and no CMU-c | 688 (32.9%) | 1 640 236 (35.0%) |
| **Localized potential access of the commune of residence / n (%)** | | |
| >4 consultations/year/inhabitant | 1 116 (53.4%) | 2 347 727 (50.1%) |
| 2.5-4 consultations/year/inhabitant | 802 (38.4%) | 1 878 938 (40.1%) |
| <2.5 consultations/year/inhabitants | 154 (7.4%) | 389 057 (8.3%) |
| **Comorbidities** | | |
| **Psychatric / n (%)** | 224 (10.7%) | 113 654 (2.4%) |
| **Hypertension / n (%)** | 54 (2.6%) | 66 077 (1.4%) |
| **Obesity / n (%)** | 96 (4.6%) | 114 974 (2.5%) |

| | | |
|---|---|---|
| **Maternal diabetes / n (%)** | | |
| No diabetes | 1 826 (87.3%) | 4 268 929 (91.1%) |
| Gestational diabetes | 234 (11.2%) | 391 412 (8.4%) |
| Pregestational diabetes | 31 (1.5%) | 23 403 (0.5%) |
| **Lifestyle** | | |
| **Tobacco use / n (%)** | 170 (8.1%) | 242 123 (5.2%) |
| **Opioids / n (%)** | 34 (1.6%) | 14 280 (0.3%) |
| **Alcohol consumption / n (%)** | 53 (2.6%) | 27 915 (0.6%) |
| **Drug exposure** | | |
| **Folic acid / n (%)** | 1 378 (65.9%) | 1 538 097 (32.8%) |
| **Psychotropic drugs / n (%)** | 159 (7.6%) | 114 997 (2.5%) |

**Table S9- Sensitivity Analyses of Association between Valproate Exposure and Major Congenital Malformations algorithm in Live births (2010 - 2015)**

| | Exposed to valproate n (n per 10.000 live births) | Non-exposed to AE drugs n (n per 10.000 live births) | crude OR [95%-CI] | adjusted OR [95%-CI] |
|---|---|---|---|---|
| Any MCM / identified with a more sensitive algorithm | 138 (660.0) | 107,727 (230.0) | 3.0 [2.5 - 3.6] | 2.8 [2.6 - 3.3] |
| Alternative endpoint: Pregnancy interruption after 22 weeks of amenorhea, still-birth or live birth with any anomaly | 180 (858.0) | 135,043 (290.0) | 3.1 [2.7 - 3.6] | 2.8 [2.4 - 3.3] |

*Figure S1 – Yearly prevalence of MCMs by organ group in EPI-MERES and EUROCAT*

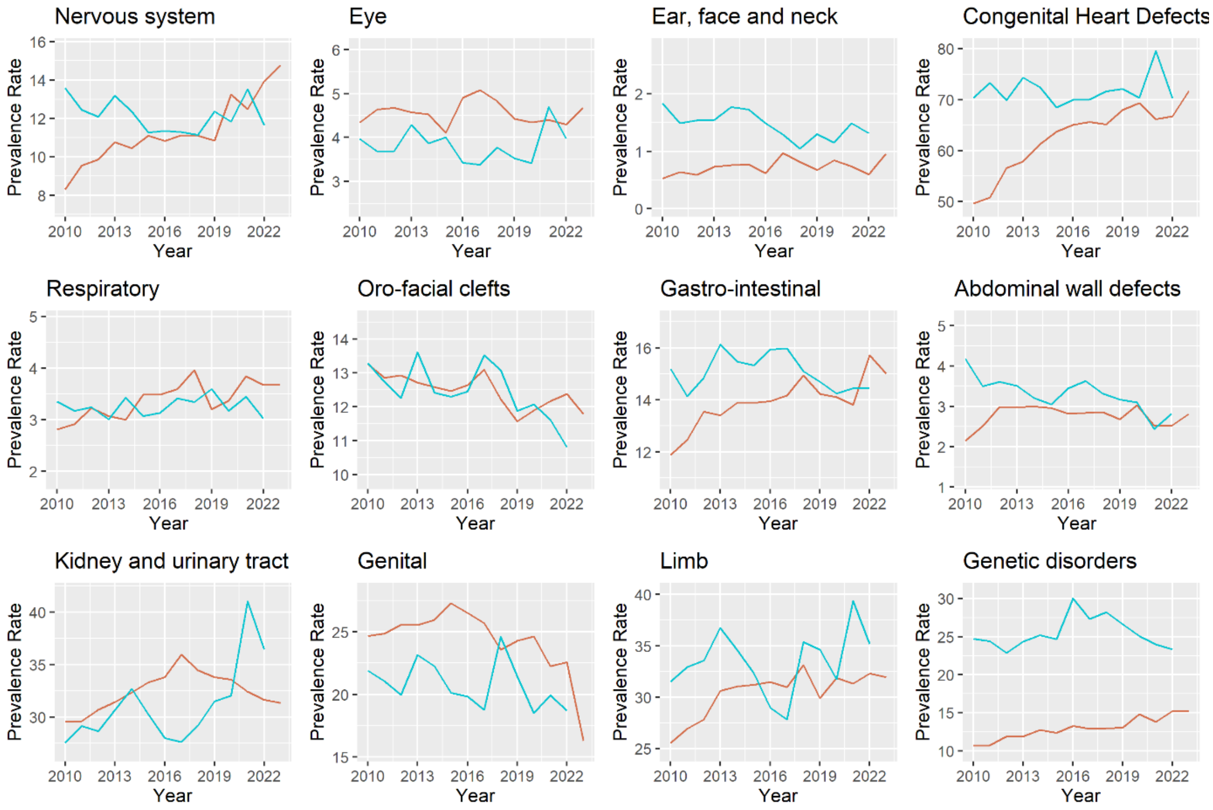

*Figure S2 – Flowchart of the valproate study*

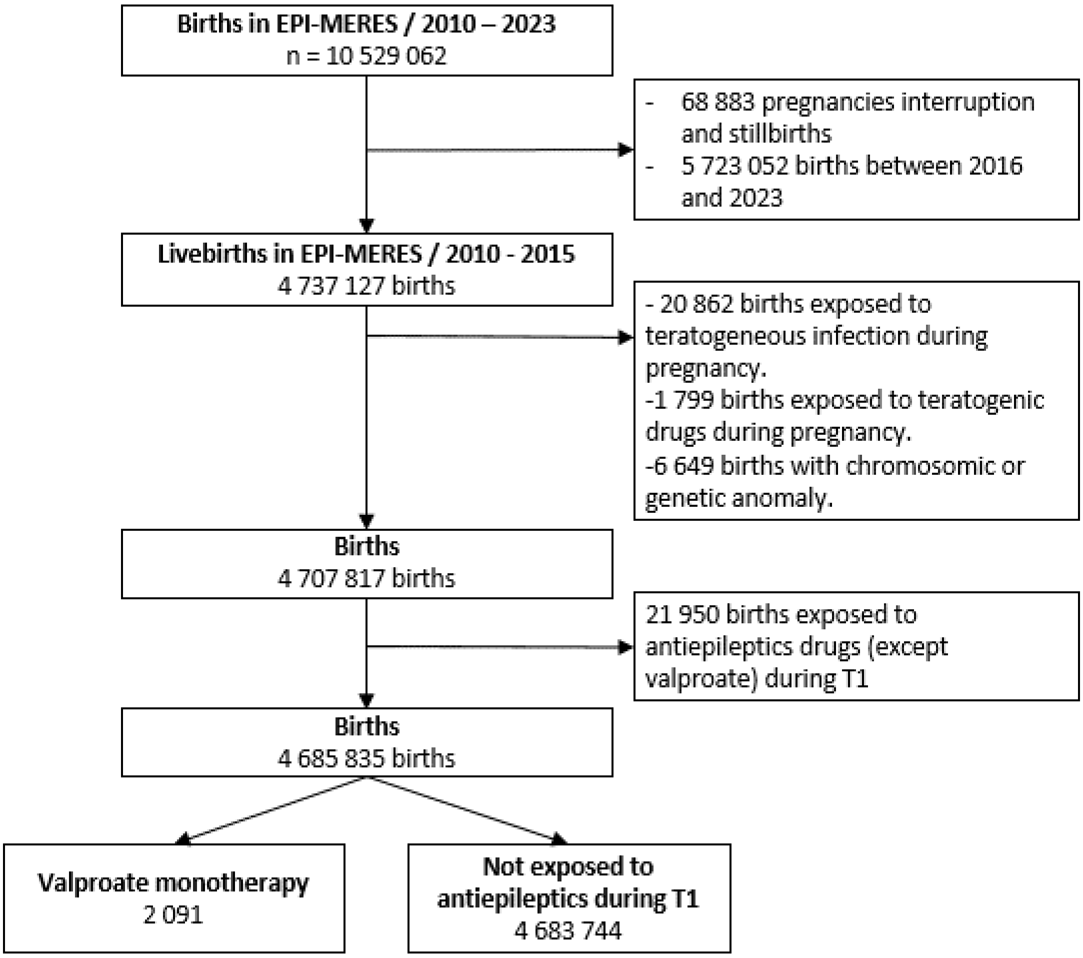

***Figure S3 – Standardized mean differences of covariates before and after propensity score matching for valproate vs no-exposure***

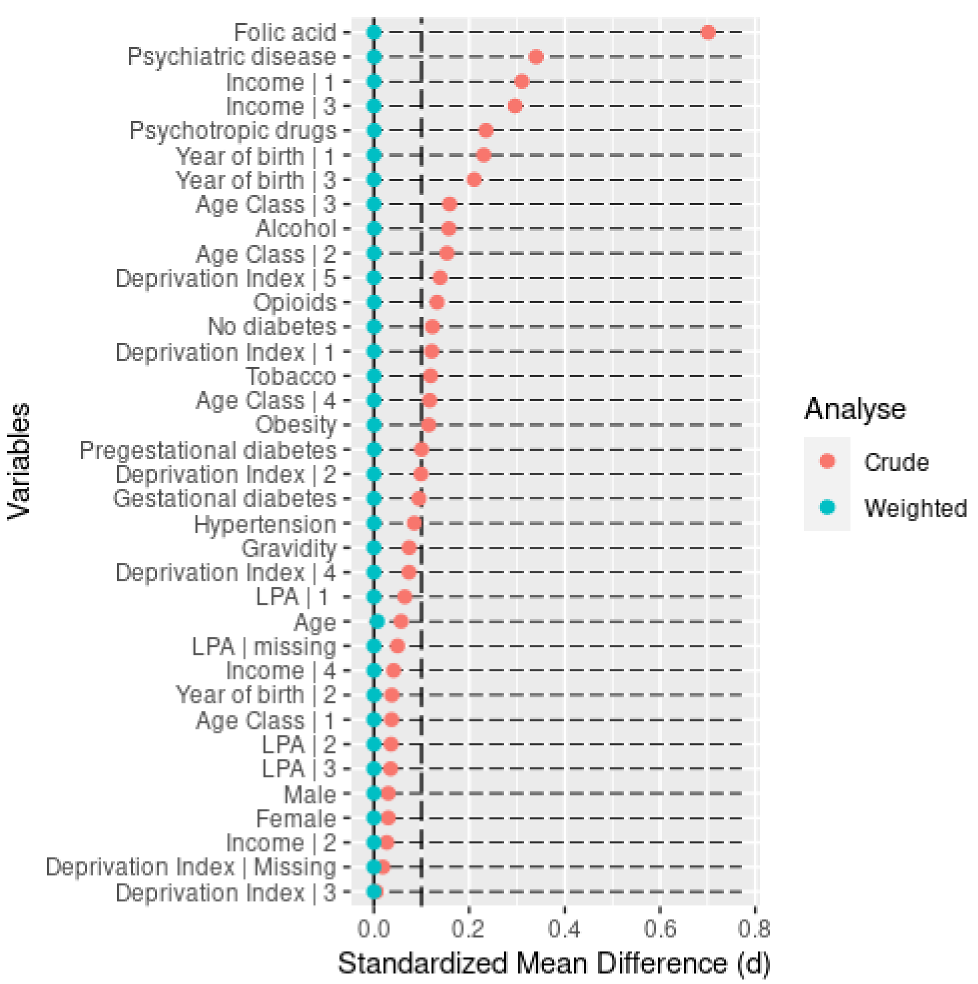
